# Comparison of cardiovascular health profiles across population surveys from five high- to low-income countries

**DOI:** 10.1101/2023.07.26.23293185

**Authors:** Lisa Ware, Bridget Vermeulen, Innocent Maposa, David Flood, Luisa CC Brant, Shweta Khandelwal, Kavita Singh, Sara Soares, Neusa Jessen, Gastón Perman, Baizid Khoorshid Riaz, Harshpal Singh Sachdev, Norrina B Allen, Darwin R Labarthe

## Abstract

**Aims:** With the greatest burden of cardiovascular disease morbidity and mortality increasingly observed in lower-income countries least prepared for this epidemic, focus is widening from risk factor management alone to primordial prevention to maintain high levels of cardiovascular health (CVH) across the life course. To facilitate this, the American Heart Association (AHA) developed CVH scoring guidelines to evaluate and track CVH. We aimed to compare the prevalence and trajectories of high CVH across the life course using nationally representative adult CVH data from five diverse high- to low-income countries.

**Methods:** Surveys with CVH variables (physical activity, cigarette smoking, body mass, blood pressure, blood glucose, and total cholesterol levels) were identified in Ethiopia, Bangladesh, Brazil, England, and the United States (US). Participants were included if they were 18-69y, not pregnant, and had data for these CVH metrics. Comparable data were harmonized and each of the CVH metrics was scored using AHA guidelines as high (2), moderate (1), or low (0) to create total CVH scores with higher scores representing better CVH. High CVH prevalence by age was compared creating country CVH trajectories.

**Results:** The analysis included 28,092 adults (Ethiopia n=7686, 55.2% male; Bangladesh n=6731, 48.4% male; Brazil n=7241, 47.9 % male; England n=2691, 49.5% male, and the US n=3743, 50.3% male). As country income level increased, prevalence of high CVH decreased (>90% in Ethiopia, >68% in Bangladesh and under 65% in the remaining countries). This pattern remained using either five or all six CVH metrics and following exclusion of underweight participants. While a decline in CVH with age was observed for all countries, higher income countries showed lower prevalence of high CVH already by age 18y. Excess body weight appeared the main driver of poor CVH in higher income countries, while current smoking was highest in Bangladesh.

**Conclusion:** Harmonization of nationally representative survey data on CVH trajectories with age in 5 highly diverse countries supports our hypothesis that CVH decline with age may be universal. Interventions to promote and preserve high CVH throughout the life course are needed in all populations, tailored to country-specific time courses of the decline. In countries where CVH remains relatively high, protection of whole societies from risk factor epidemics may still be feasible.

## Introduction

Globally, cardiovascular diseases (CVD) remain the leading cause of mortality and an important contributor to morbidity.^1^ In terms of years of life lost, diminished quality of life, as well as direct and indirect medical costs, the burden of CVD is enormous.^2^ As a result of ongoing epidemiological transitions, almost 80% of global CVD related deaths occur in low- and middle-income countries (LMICs).^1^ As health systems are overwhelmed, there are increasing shifts to primary prevention (risk factor modification) and ultimately primordial prevention (avoidance of risk factor onset) to maintain good cardiovascular health (CVH) for as long as possible and reduce the burden of CVD.^2^

In 2010, the American Heart Association (AHA) defined ideal CVH as a combination of seven metrics - Life’s Simple 7 (LS7: dietary quality, physical activity (PA), exposure to cigarette smoke, body mass index (BMI), blood pressure (BP), blood glucose, and total cholesterol). All metrics could be characterized as either ideal (2 points), intermediate (1 point) or poor (0 point) to generate a combined CVH score from 0 to 14 points and good CVH defined as having 5 – 7 metrics at an ideal level.^2^ In June 2020, the AHA introduced an updated CVH score - Life’s Essential 8 (LE8) which revised many of the components of LS7 as well as adding a new metric (sleep) and a revised scoring system (with scores 0 to 100).^3^

Previous studies have shown that good CVH strongly associates with lower risk of CVD morbidity and mortality, and all-cause mortality in young, middle-aged, and older adults.^4–10^ However, the prevalence of good CVH typically decreases with age and by adulthood is generally low, though evidence from LMICs is lacking.^10, 11^ To better understand the differences in CVD burden between HICs and LMICs, population level data are needed that capture the CVH metrics of interest including health behaviors and sociodemographic profiles. Large population health surveys, as opposed to clinical data, have the potential to inform countries’ health planning and promotion strategies and understanding of the future burden of poor CVH, while identifying subgroups at particular risk.^12^

A nationally representative, population-based survey in Venezuela showed that two-thirds of the population had poor or only intermediate CVH,^13^ while an analysis of population CVH using the Brazilian Health Survey suggested over half (56.7%) of the population had ideal CVH.^14^ However, differences in the use and application of the CVH score in these neighboring countries obscures country comparisons. Additionally, much of the prior research investigating CVH has originated from HICs including the United States (US) where less than 20% of adults are reported to have ideal CVH.^15^ Therefore, this research sought to explore the availability and comparability of nationally representative adult CVH data from five high- to low-income countries and to compare the CVH prevalence and patterns of decline. The aim was to test the hypothesis that the decline in CVH with age is universal, though this may differ in starting level and rate of change potentially revealing influential factors and inflection points that may be discernible as points of intervention.

## Methods

### Data sources and study design

We purposely selected five countries that were diverse along dimensions of geographic location, per-capita income, burden of disease, educational systems, and health system capacity. Population surveys were assessed for the presence of the eight CVH metrics of interest, identifying the World Health Organization (WHO) STEPwise approach to surveillance (STEPS) surveys as an important source of data covering over 120 countries from across all WHO world regions.^16^ Datasets from Bangladesh and Ethiopia were selected for analysis as low and lower-middle income countries from Africa and South Asia.

The suitability of WHO-STEPS for CVH assessment is perhaps unsurprising as it was designed as a standardized non-communicable disease (NCD) risk surveillance tool with a manual for collecting, analyzing and disseminating data.^17^ As such, WHO-STEPS data are designed to be directly comparable between implementing countries, many of which are lower income countries. To undertake a comparison with upper-middle- and high-income country data, the US National Health and Nutrition Examination Survey (NHANES), the Health Survey for England (HSE) and the Brazilian National Health Survey (PNS) were identified as nationally representative surveys with CVH data available. These five countries span a range from low to high income as defined by the World Bank,^18^ with the UK and US as HICs, Brazil as an upper middle-income country, Bangladesh as a lower middle-income country, and Ethiopia as a low income country.

Permission was requested from respective data owners and access was granted to cross-sectional survey data from these five countries: WHO-STEPS surveys from Bangladesh (2018) and Ethiopia (2015);^16, 17, 19^ US NHANES pre-pandemic data 2017-2020;^20^ HSE 2016;^21^ and PNS-Brazil 2013.^14 22^ These five countries were also selected as examples with varying structural systems that may influence lifetime CVH. For example, Bangladesh has only five years of compulsory education, Ethiopia has eight years, the UK has 11 years, the US has 12 years, and Brazil has 14 years.^23^ Health expenditure also varies from $71 per capita in Ethiopia to almost $11,000 per capita in the US.^24^ The causes of mortality in the five countries also vary, with the percentage of deaths from maternal and child health, nutrition or communicable disease at 45% of deaths in Ethiopia and 5% in the US.^25^

All surveys obtained ethical permission from relevant national/institutional ethics committees before implementation. For this specific secondary data analysis, ethical approval was granted from the University of the Witwatersrand Medical Human Research Ethics Committee [Ref: M220437].

### Surveys and Participants

In combination, the five selected surveys included 49,774 participants: STEPS Bangladesh (2018)^26^ - 8185 adults (aged 18 – 69 years); STEPS Ethiopia 2015^19^ - 9251 adolescents and adults (aged 15 – 69 years); NHANES (US) 2017-2020^20^ - 15 560 children and adults (aged 0 – 80 years); HSE (England) 2016^27^ - 7826 children and adults (aged 0 – 90 years); and PNS-Brazil 2013^22^ - 8952 adults (aged 18 – 104 years). For both NHANES and HSE, data collection occurred in stages. For HSE, there was a home interview and a nurse visit, while participants in NHANES undertook home interviews, health assessments at a mobile examination center (MEC), clinical examinations, and laboratory investigations. Not all participants are selected for or complete all stages. Only those participants with survey, health and clinical exam, and lab analysis (as evidenced by data on total cholesterol, HbA1c, BMI, BP, physical activity, smoking) from NHANES were included in this analysis. Only HSE participants with home interview and nurse visit who agreed to give blood were included. Participants who were pregnant, or aged under 18 or over 69 years, or with missing data were excluded.

### Cardiovascular health data and harmonization

Availability of CVH variables (dietary quality, sleep, PA, cigarette smoking, BMI, BP, blood glucose or HbA1c, and total cholesterol levels) and the methods of data collection were compared across surveys to make decisions regarding harmonization (**Figure 1**). The data collection methods for each of the CVH metrics are reviewed briefly in Suppl. Table 1 with the full methodology for each survey described elsewhere.^14, 16 17 19–22, 26–29^ Data that were considered sufficiently comparable were harmonized (**Suppl. Table 1**). Harmonized variables were also created for education and current medication use prescribed to reduce BP, glucose, or cholesterol (**Suppl. Figure 1**), with age and sex data extracted from each survey.

**Figure 1.**
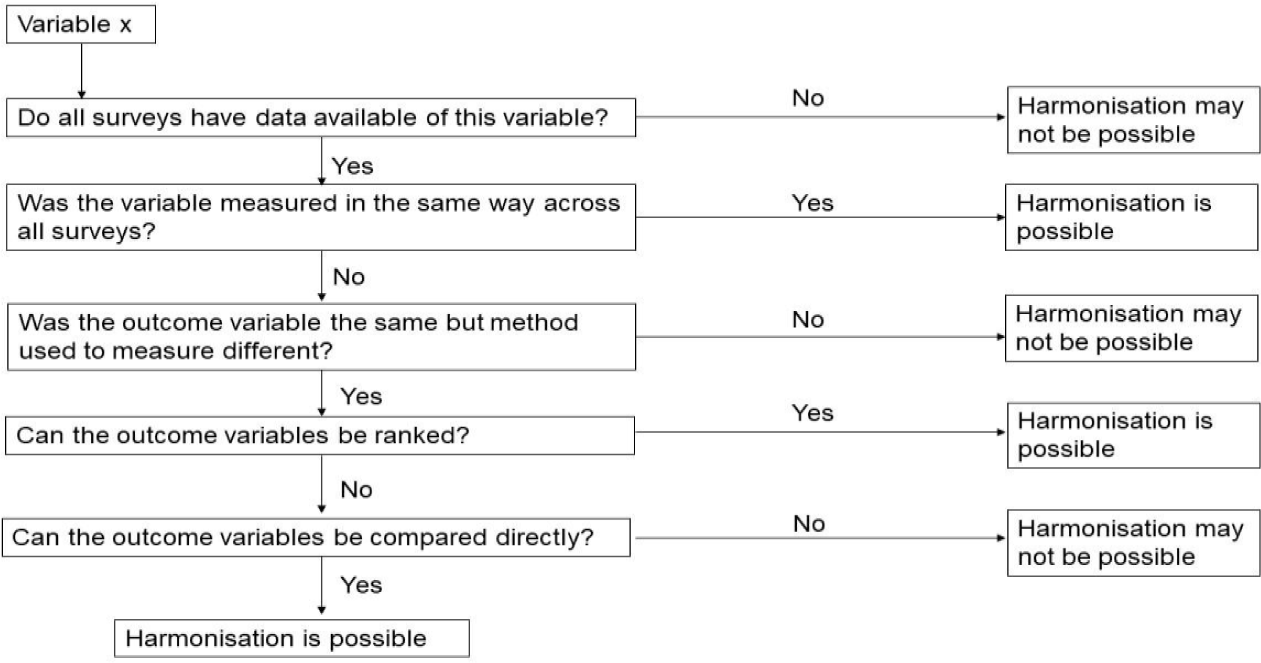
Harmonization decision matrix (adapted from Schaap et al.^30^)

### Data analysis

The data available on dietary quality and sleep could not be harmonized. The six remaining CVH metrics (PA, cigarette smoking, BMI, BP, blood glucose/HbA1c, and total cholesterol levels) were harmonized and categorized as shown in **Table 1** (and Suppl. Table 1) based on both the LS7 and LE8 guidelines. ^2, 3^ This secondary data analysis was conceived under the LS7 paradigm, however, during the process of analysis the LE8 criteria were published and therefore we utilized elements of both LS7 and LE8 that best fit with our datasets. A score of 2 was allocated for each CVH metric in the high category, 1 for moderate and 0 for low, with scores summed to create a combined CVH score categorized as follows: for six CVH metrics, 9-12 was high CVH, 6-8 was moderate and <6 was low; for five CVH metrics, 8-10 was high CVH, 5-7 was moderate and <5 was low. Statistical analyses were performed with IBM^®^ SPSS^®^ Statistics, Version 28 (IBM Corporation, Armonk, New York). Participant characteristics were described by country with results reported as number of participants (n) or weighted percentage (%) using weights developed by each of the surveys as described elsewhere.^19, 26, 31^ Normality of data was determined with visual inspection of histograms. The prevalence of high CVH was determined for each country, comparing categorical variables using Chi-square tests and comparing weighted prevalence (arithmetic mean ± standard deviation) of each CVH metric across countries using ANNOVA. To create CVH trajectories, age was categorized into five-year bands and the median of each age band was plotted against the weighted age-specific prevalence of high CVH.

**Table 1.** Cardiovascular health (CVH) scores for adults 18-69 years based on Lloyd-Jones et al.^2, 3^.

|  | Cardiovascular (CVH) health scores |  |  |
| --- | --- | --- | --- |
| CVH Metric | LS7 | LE8<sup>a</sup> | Adapted combined |
| *Body mass index (kg/m<sup>2</sup>)* |  |  |  |
| High CVH | <25 | <25 | As LS7 & LE8 |
| Moderate CVH | 25–29.9 | 25–29.9 | As LS7 & LE8 |
| Low CVH | ≥30 | ≥30 | As LS7 & LE8 |
| *Blood pressure (mmHg)* |  |  |  |
| High CVH | <120/80 (-meds) | <120/80 (+/-meds) | As LS7 |
| Moderate CVH | 120–139/80–89 (-meds) or treated to goal | 120–129/<80 (+/-meds) or 130–139/80–89 (-meds) | 120–139/80–89 (-meds) or <140/90 (+meds) |
| Low CVH | ≥140/90 (+/-meds) | 130–139/80–89 (+meds) or ≥140/90 (+/-meds) | As LS7 |
| *Cholesterol level (mg/dL) (Blood lipids)* |  |  |  |
| High CVH | Total cholesterol <200 (-meds) | Non-HDL cholesterol <130 (+/-meds) | As LS7 |
| Moderate CVH | 200–239 (-meds) or treated to goal | 130–159 (-meds) | 200–239 (-meds) or <240 (+meds) |
| Low CVH | ≥240 | 130–159 (+meds) or ≥160 (+/-meds) | ≥240 (+/-meds) |
| *Fasting blood glucose (mg/dL)* |  |  |  |
| High CVH | <100 (-meds) | <100 and no history of diabetes | As LS7 |
| Moderate CVH | 100–125 (-meds) or | 100–125 and no diabetes | 100–125 (-meds) or |
|  | treated to goal |  | <126 (+meds) |
| Low CVH | ≥126 | ≥126 or Diabetes | ≥126 (+/-meds) |
| *HbA1c (%)* |  |  |  |
| High CVH | NA | No history of diabetes and <5.7 | <5.7 (-meds) |
| Moderate CVH | NA | No diabetes and 5.7–6.4 | 5.7-6.4 (-meds) or <6.5 (+meds)* |
| Low CVH | NA | ≥6.5 or diabetes | ≥6.5 (+/- meds) |
| *Nicotine exposure (self-reported use)* |  |  |  |
| High CVH | Never smoked or quit >12 months | Never used combustible tobacco or inhaled nicotine delivery systems (NDS) | As LS7 |
| Moderate CVH | Former ≤12 months | Former ≥1 year or if living with active indoor smoker at home, former ≥5 years | As LS7 |
| Low CVH | Current smoker | Current combustible tobacco or NDS use or former <1 year | As LS7 |
| *Physical activity (self-reported minutes/week)* |  |  |  |
| High CVH | ≥150 MVPA or ≥75 VPA | ≥90 MVPA | As LS7 |
| Moderate CVH | 1-149 MVPA or 1-74 VPA | 60–89 MVPA | As LS7 |
| Low CVH | None | <60 MVPA | As LS7 |
| *Healthy diet (self-reported)* |  |  |  |
| High CVH | 4-5 components | ≥75th percentile (populations) or MEPA score 12-16 (individuals) | Not included |
| Moderate CVH | 2-3 components | 50 – 74th percentile or MEPA score 8-11 | Not included |
| Low CVH | 0-1 components | <50th percentile or MEPA<8 | Not included |
| *Sleep health (self-reported average hours of sleep per night)* |  |  |  |
| High CVH | Not included | 7 – <10 hours | Not included |
| Moderate CVH | Not included | 6 – <7 hours | Not included |
| Low CVH | Not included | <6 or ≥10 hours | Not included |

Abbreviations: MVPA, moderate and vigorous physical activity; MPA, moderate physical activity; VPA, vigorous physical activity; Meds, medicines; MEPA, Mediterranean Eating Pattern for American questionnaires; NA, not applicable; NDS, nicotine-delivery system. <sup>a</sup>LE8 is scored on a 0-100 scale with CVH scores of 80 to 100 considered high CVH; 50 to 79, moderate CVH; and 0 to 49 points, low CVH. \*Glycaemia assessed using either fasting glucose or HbA1c. Moderate HbA1c scoring applied to include treated to goal to align with analysis of fasting blood glucose data.

## Results

Among the 49,774 participants (0-104 years) across the surveys, 28,092 adults (18 – 69 years) met the inclusion criteria for this secondary data analysis (**Figure 2**). Exclusion by age ranged from zero percent in the STEPS surveys, to 21% of the survey population in HSE, while exclusion for pregnancy was 4% or less across all surveys. The most frequently observed missing data in the WHO-STEPS and PNS surveys were the glycaemia and cholesterol measures (Bangladesh 14% and 14% missing; Ethiopia 10% and 9% missing; Brazil 4% and 4% missing). For BMI and BP, the percentage of cases with missing data were Bangladesh 2% and 0.3%; Ethiopia 5% and 1%; Brazil 1% and 1%. For smoking and PA, the percentage of cases with missing data were generally low (Bangladesh 0% and 2%; Ethiopia 0.1% and 3%; Brazil 0.1% and 1%). Following exclusion of participants based on age and/or pregnancy, the exclusion of cases due to missing data ranged from 8% of the survey sample in Brazil to 44% in the US. Missing data in the US and England were primarily due to missing information on current treatment use for BP, cholesterol or glycaemia and/or missing sociodemographic data.

**Figure 2.**
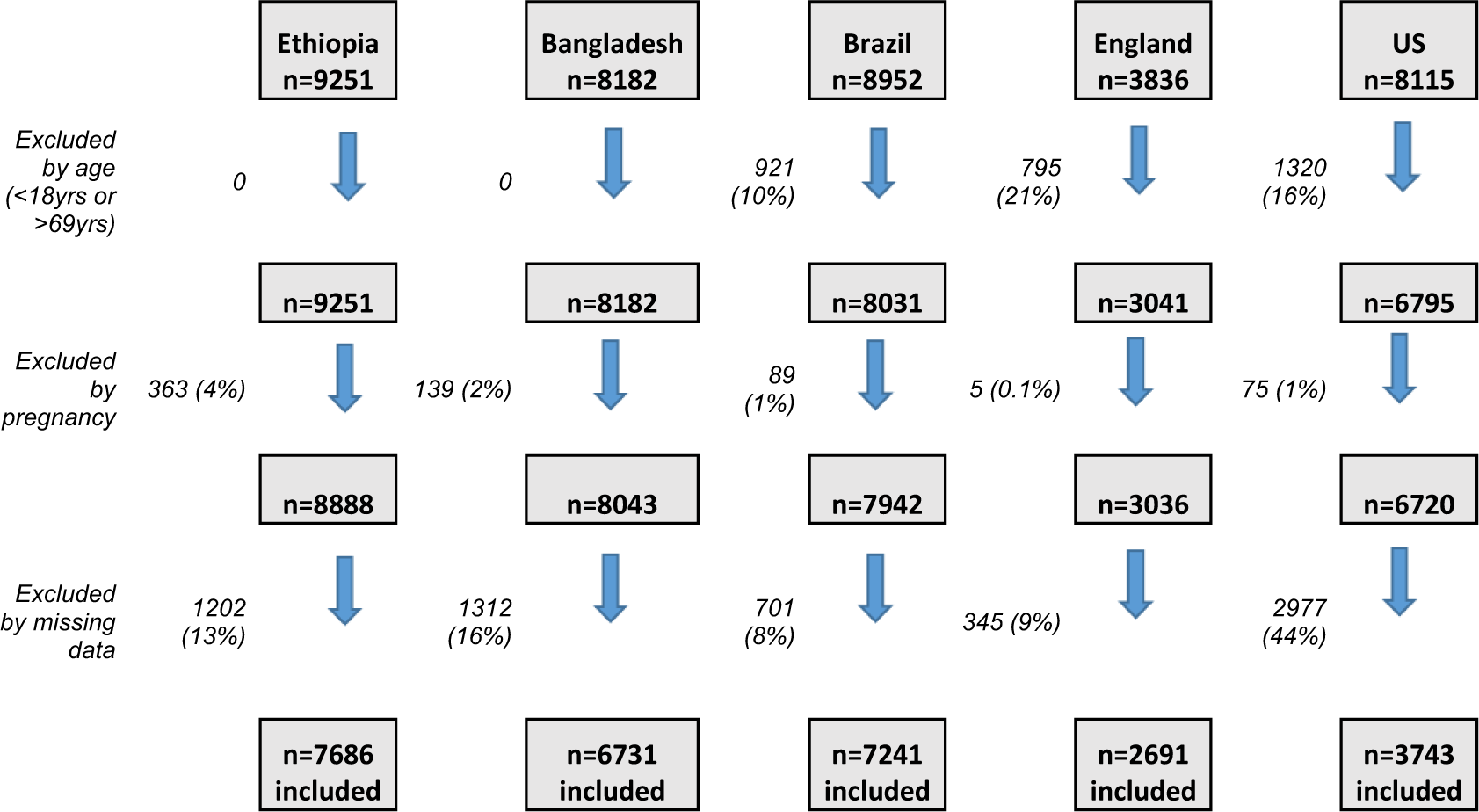
Consort diagram of participants included in the analysis.

Included participants for each country were: Ethiopia n=7686 (55.2% male); Bangladesh n=6731 (48.4% male); Brazil n=7241 (47.9 % male); England n=2691 (49.5% male), and the US n=3743 (50.3% male) (**Table 2**). Education level varied significantly between LMICs and HICs, with 88% of participants having primary school or less in Ethiopia, compared to just 3% at the same educational level in the US. The age distribution of the surveys also varied with more younger adults in the lower than in the higher income countries (% of sample 18-35y: Ethiopia 60%; Bangladesh 48.7%; Brazil 33.5%; England 25.0%; US 29.6%). There were significant differences in the population levels of all weighted mean CVH metrics between countries. The highest levels of reported physical activity were observed in Ethiopia. Including those both on treatment and with no treatment, the lowest SBP was observed in the US, the lowest DBP was observed in England, the lowest BMI, cholesterol, and glucose levels in Ethiopia, and the lowest HbA1c in Brazil. Medication use for BP, glycaemia and cholesterol was typically higher in England, US and Brazil than in Ethiopia and Bangladesh, although the prevalence of treatment use for hypertension was lower in England compared to both Brazil and the US. Treatment levels for hyperlipidemia and for diabetes generally tracked with the mean population levels of these metrics i.e., treatment increased as population mean levels increased. The exceptions to this were the US which showed the second highest mean total cholesterol level but the highest percentage on treatment for lipids (more than twice that of England), and Brazil which showed the lowest mean HbA1c levels and the highest prevalence of treatment for diabetes (twice that of England and the US).

**Table 2:** Characteristics of the adult participants included in the analysis for each country.

|  | **Ethiopia**<br>**(n=7686)** | **Bangladesh**<br>**(n=6731)** | **Brazil**<br>**(n=7241)** | **England**<br>**(n=2691)** | **United States**<br>**(n=3743)** | ***P value*** |
| --- | --- | --- | --- | --- | --- | --- |
| Age (years), range | 18 – 69 | 18 – 69 | 18 – 69 | 18 – 69 | 18 – 69 | <0.001 |
| 18-24, n (%) | 1388 (26.9) | 717 (20.7) | 540 (9.20) | 153 (8.40) | 428 (11.5) |  |
| 25-34, n (%) | 2442 (33.1) | 1739 (28.0) | 1502 (24.3) | 408 (16.6) | 577 (18.1) |  |
| 35-44, n (%) | 1794 (18.8) | 1904 (18.9) | 1806 (22.7) | 523 (20.5) | 612 (17.3) |  |
| 45-54, n (%) | 1119 (12.2) | 1356 (13.6) | 1626 (21.0) | 614 (22.7) | 728 (19.6) |  |
| 55-64, n (%) | 654 (6.60) | 812 (13.4) | 1268 (16.7) | 669 (21.6) | 985 (23.8) |  |
| 65-69, n (%) | 298 (2.60) | 203 (5.30) | 499 (6.10) | 324 (10.2) | 413 (9.6) |  |
| Male sex, n (%) | 3219 (55.2) | 3073 (48.4) | 3042 (47.9) | 1194 (49.5) | 1826 (50.3) | <0.001 |
| Highest Education level |  |  |  |  |  | <0.001 |
| Primary school or less, n (%) | 6791 (88.4) | 5282 (78.0) | 3737 (45.9) | NA | 245 (3.00) |  |
| Secondary/High school, n (%) | 493 (6.90) | 1050 (17.3) | 2370 (36.1) | 610 (21.4) | 1205 (32.1) |  |
| College/University, n (%) | 7 (0.10) | 399 (4.70) | 1134 (18.0) | 1699 (65.3) | 2143 (62.3) |  |
| Body Mass Index (kg/m<sup>2</sup>) | 20.4 ± 2.97<br>*a; e; f; g* | 22.5 ± 4.35<br>*a; b; c; d* | 26.5 ± 5.15<br>*d; g; i; j* | 27.1 ± 5.29<br>*c; f; h; j* | 30.0 ± 7.54<br>*b; e; h; i* | <0.001 |
| Underweight | 1775 (23.5) | 909 (14.8) | 169 (2.4) | 33 (1.4) | 56 (1.6) |  |
| Healthy weight | 5120 (70.0) | 3855 (60.1) | 2818 (40.1) | 955 (37.3) | 871 (24.5) |  |
| Overweight / obese | 791 (6.5) | 1967 (25.1) | 4254 (57.5) | 1703 (61.3) | 2816 (73.4) |  |
| Systolic blood pressure (mmHg) | 121 ± 17.94<br>*a; e; f; g* | 121 ± 18.29<br>*a; b; c; d* | 124 ± 17.85<br>*d; g; i* | 124 ± 15.12<br>*c; f; h* | 119 ± 15.95<br>*b; e; h; i* | <0.001 |
| Diastolic blood pressure (mmHg) | 78 ± 11.70<br>*a; e; f* | 78 ± 11.78<br>*a; b; c* | 78 ± 11.36<br>*i; j* | 73 ± 10.57<br>*c; f; j* | 74 ± 10.84<br>*b; e; i* | <0.001 |
| On treatment for BP, n (%) | 120 (1.1) | 545 (6.0) | 1125 (15.1) | 303 (9.5) | 1030 (23.1) | <0.001 |
| Total cholesterol (mg/dL) | 131 ± 31.95<br>*a; e; f; g* | 170 ± 37.48<br>*a; b; c; d* | 184 ± 38.62<br>*d; g; j* | 198 ± 41.68<br>*c; f; h; j* | 186 ± 41.48<br>*b; e; h; i* | <0.001 |
| On treatment for lipids, n (%) | 8 (0.0) | 76 (0.8) | 648 (3.7) | 279 (8.2) | 738 (18.2) | <0.001 |
| Glucose (mg/dL) | 79.6 ± 20.24<br>*a; e; f; g* | 97.5 ± 31.67<br>*a; b; c; d* | NA | NA | NA | <0.001 |
| Glycated hemoglobin (HbA1c %) | NA | NA | 5.47 ± 0.95<br>*d; g* | 5.53 ± 0.76<br>*c; f* | 5.76 ± 1.10<br>*b; e* | <0.001 |
| On treatment for diabetes, n (%) | 51 (0.5) | 243 (2.9) | 284 (8.1) | 111 (4.1) | 191 (3.9) | <0.001 |
| Total active minutes per week | 2111 ± 1839<br>*a; e; f; g* | 1462 ± 1983<br>*a; b; c; d* | 71 ± 174<br>*d; g; i; j* | 521 ± 688<br>*c; f; h; j* | 897 ± 1232<br>*b; e; h; i* | <0.001 |

Abbreviations: NA, data not available; BP, blood pressure. All values presented are means and standard deviations (SD) unless otherwise indicated. Weighted values are reported for mean ± SD and for percentages. For smoking prevalence, see Table 4 (poor = current smoker, intermediate = former smoker, good = never smoker). Letters in *italics* denote statistical significance at $p < 0.001$ between the groups sharing the same letter.

The mean CVH score for each country and the prevalence of high CVH (for all six metrics combined and for each metric) are shown in **Table 3**. Based on the combination of all six metrics, Brazil had the lowest mean CVH score (7.7/12) with 38.7% of the population scoring as high CVH and Ethiopia had the highest (10.5/12) with 91.2% of the population scoring as high CVH. Examining each of the CVH metrics, there were clear patterns between countries in income level, BMI, and cholesterol, such that as country income level increased, both BMI and cholesterol scores decreased. Glycaemia showed a similar trend with less prevalent low glycaemia scores (≥126 mg/dl fasting blood glucose or ≥ 6.5% HbA1c) in the lowest income country (Ethiopia, 2%) and the highest prevalence in the US (12.6%). However, Bangladesh, England and Brazil showed similar levels, ranging from 5.2-7.1% with low scores for glycaemia.

**Table 3:**
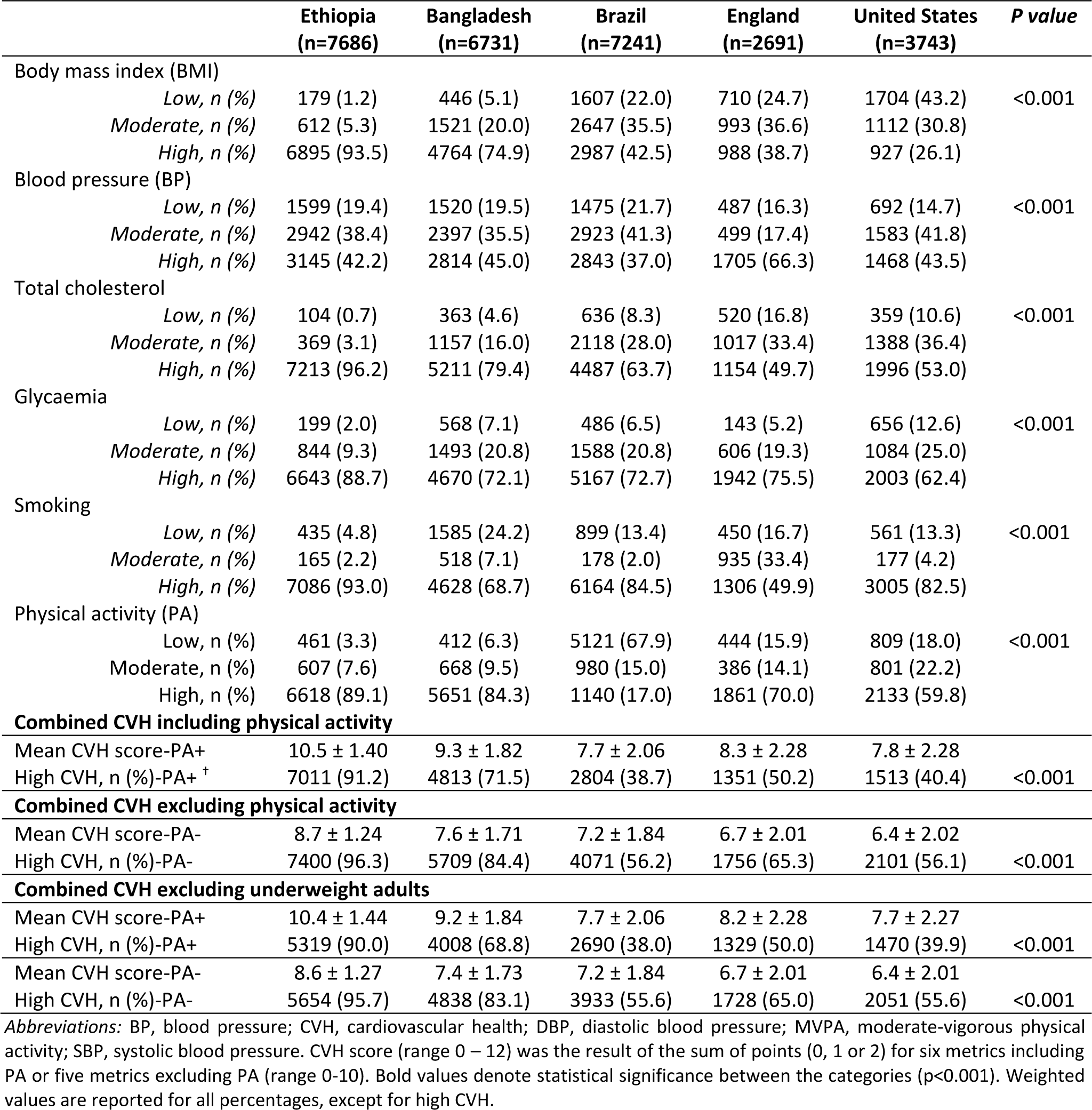
Prevalence of high, moderate, and low cardiovascular health by country study population.

Blood pressure was also more similar across the countries, with 14.7-21.7% of individuals scoring low (BP≥140/90 mmHg). Although, the pattern in England appeared different with fewer individuals in the moderate BP category (pre-hypertensive or hypertensive and treated to goal), and relatively more in the high CVH BP category (<120/80 mmHg untreated) compared to other countries. To investigate this further, we revisited the medication use for BP in England and found that, while 303 (9.5% of adults) were taking medication prescribed for BP, a higher number of adults (n=371, 11.6% of adults) reported taking medications that were known to influence BP yet not specifically prescribed for BP. As such, this may have affected the recorded BPs but would not feature in the classification of BP for the CVH score, potentially inflating the number of individuals classified as scoring high for BP. Other surveys did not have such data.

While Ethiopia, Brazil and the US had over 80% of the population who had never smoked (high scores), this percentage was lower in England (49%) and in Bangladesh (69%). In England, this was due to a high prevalence of former smokers (33.4%), while in Bangladesh this was due to a higher prevalence of current smokers (24.2%).

The prevalence of low physical activity scores ranged from 3-18% in all countries, except for Brazil (67.9%). To investigate further this large difference, we again reviewed the physical activity self-report instruments used. Further examination of the methodology revealed that the PA instrument used in Brazil only queried leisure time activity, while both the GPAQ and IPAQ used in other countries queried physical activity across multiple domains including work, leisure time and travel. Due to the risk that physical activity results were not comparable, country median CVH scores and percentage ideal CVH were recalculated excluding PA. Following the removal of the PA metric, the mean CVH scores decreased for all countries, with Ethiopia still having the highest mean score (8.7/10) but the US now moving to the lowest overall CVH scoring position (6.4/10). Additionally, the percentage of individuals classified with high CVH based on these five metrics increased for all countries to range from 56% in both the US and Brazil to 96% in Ethiopia.

Examining further the BMI data, we investigated the percentage of individuals that were scored as high CVH but that had an underweight BMI (<18.5 kg/m^2^). This was particularly high in Ethiopia with underweight individuals making up one quarter (25.7%, n=1775) of those adults scoring high for BMI (BMI <25 kg/m^2^). An analysis removing those individuals classified as underweight did not significantly affect the mean country CVH scores (Ethiopia 10.5 ± 1.40 including all adults; 10.4 ± 1.44 excluding underweight adults) or the percentage of adults scoring high for CVH (Ethiopia 91.2% including all adults; 90.0% excluding underweight adults).

The prevalence and trajectories of high CVH by age starting in adolescents (age 18y) were then plotted by country for comparison both including and excluding the PA metric for comparison (**Figures 3a and 3b**). In all countries, a decline in high CVH with aging was observed, as was our initial hypothesis. The levels of high CVH observed in adolescents also varied by country as per our initial hypothesis. When physical activity was included, levels of high CVH in adolescents were highest in Ethiopia, followed by Bangladesh, England, US and then Brazil. This ordering was maintained across the adult age span but with steeper declines observed in Brazil, the US and England compared to Bangladesh and Ethiopia such that there were marked differences in the levels of high CVH by age 69y between countries and the lower income countries appearing to perform much better than the higher income countries.

With PA excluded, Ethiopia continued to have a high starting point and a shallower decline in high CVH across the life course, although the relative advantage among adolescents appeared also in Bangladesh and the USA, and to a lesser extent England, with Brazil starting at a lower point than other countries indicating relatively fewer adolescents from Brazil entering adulthood with high CVH scores. The steepest declines in CVH were observed in the highest income countries and followed the order of country income level i.e., USA, then England and Brazil, followed by Bangladesh and lastly Ethiopia. However, the overall pattern in all countries and in both sets of analysis (**Figure 3a, 3b**) was the same showing a decreased prevalence of high CVH as age increased.

**Figure 3a.**
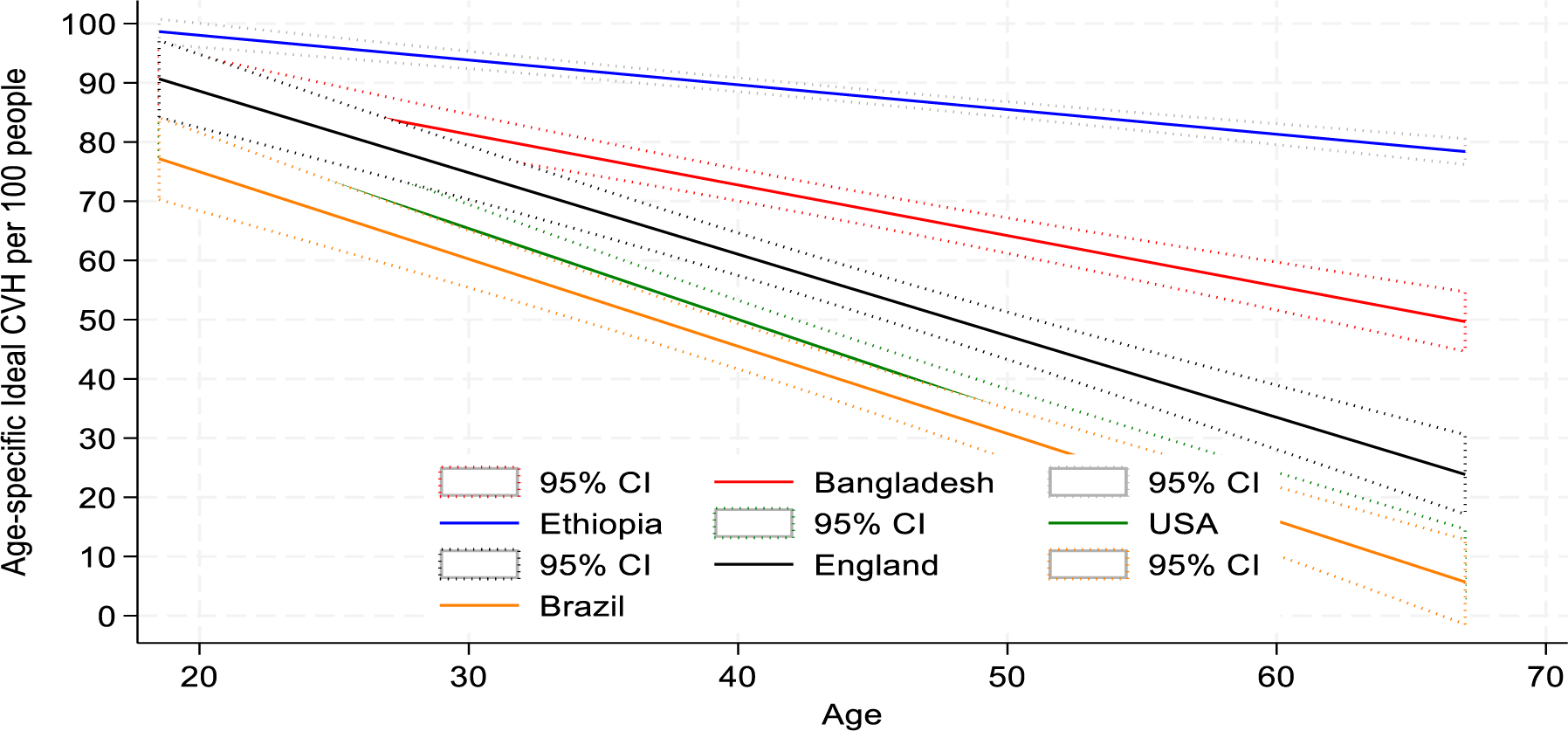
Age-specific high CVH rates for the five countries (adults aged 18-69y) – Based on six metrics (BP, BMI, cholesterol, glycaemia, smoking, PA)

**Figure 3b.**
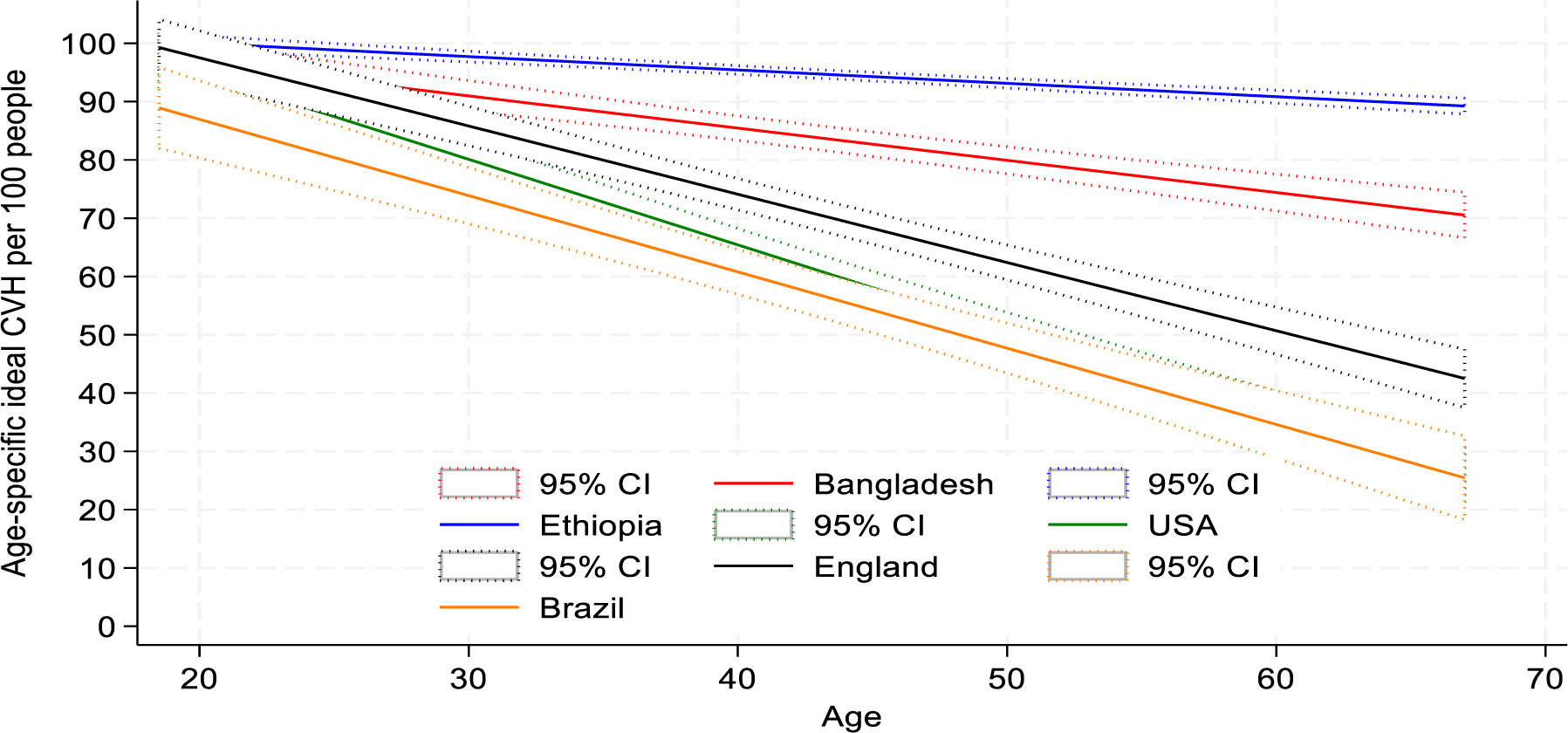
Age-specific high CVH rates for the five countries (adults aged 18-69y) – Based on five metrics (BP, BMI, cholesterol, glycaemia, smoking)

The prevalence of sex-specific CVH risk was plotted to determine whether sex specific CVH differences exist and/are consistent across higher income, lower-middle income and lower-income countries, both including and excluding the PA metric for comparison (**Figures 4a and 4b**). When analysis was conducted on all six CVH metrics (including PA), younger men presented with significantly better CVH scores in Brazil than their female counterparts (**Figure 4a**). When PA was excluded from the analysis, with the exception of Ethiopia, women generally had a better CVH than men in most countries and especially at younger ages (**Figure 4b**).

**Figure 4a.**
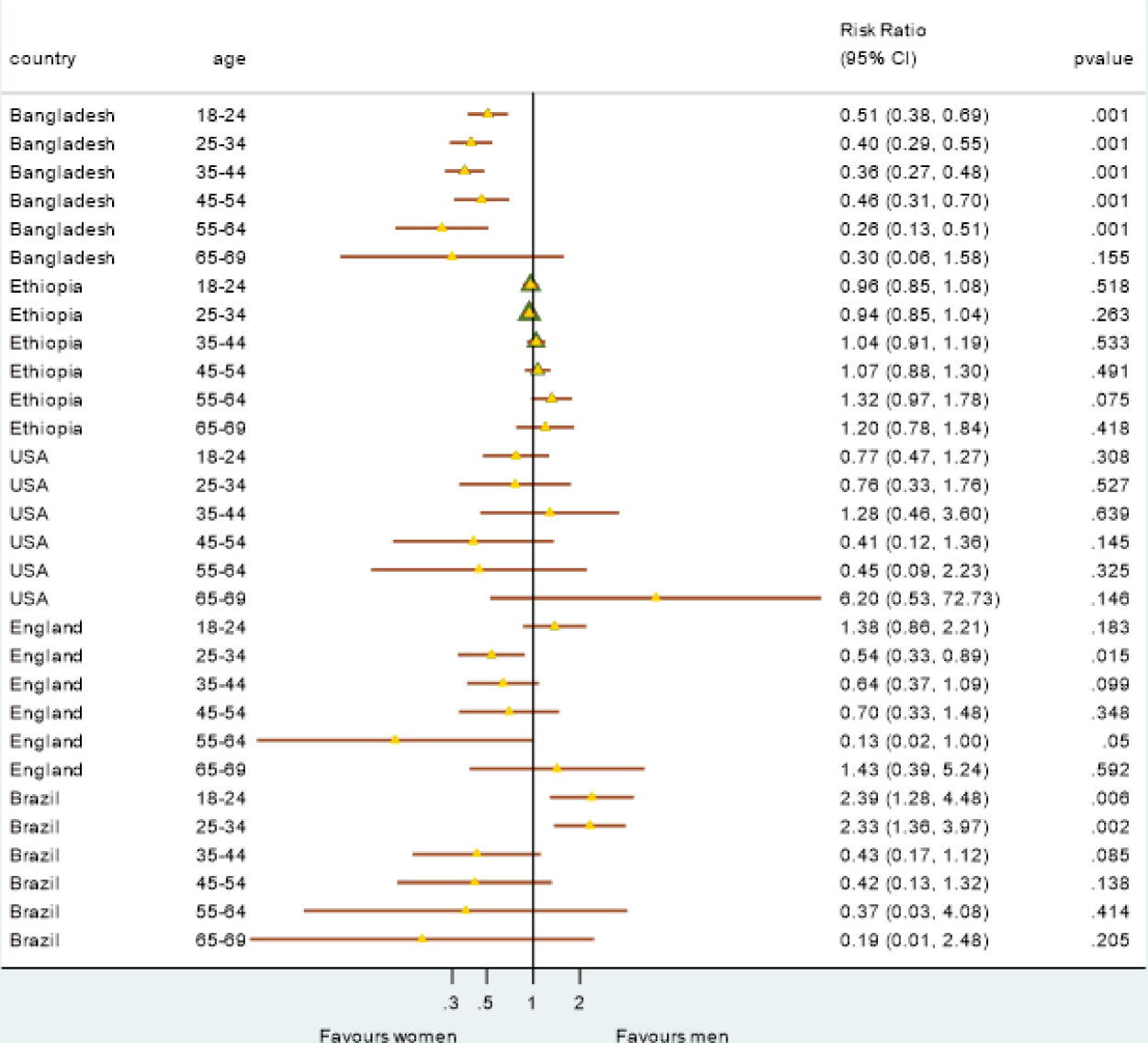
Sex specific ideal CVH risk by age group for each country - Based on six metrics (BP, BMI, cholesterol, glycaemia, smoking, PA)

**Figure 4b.**
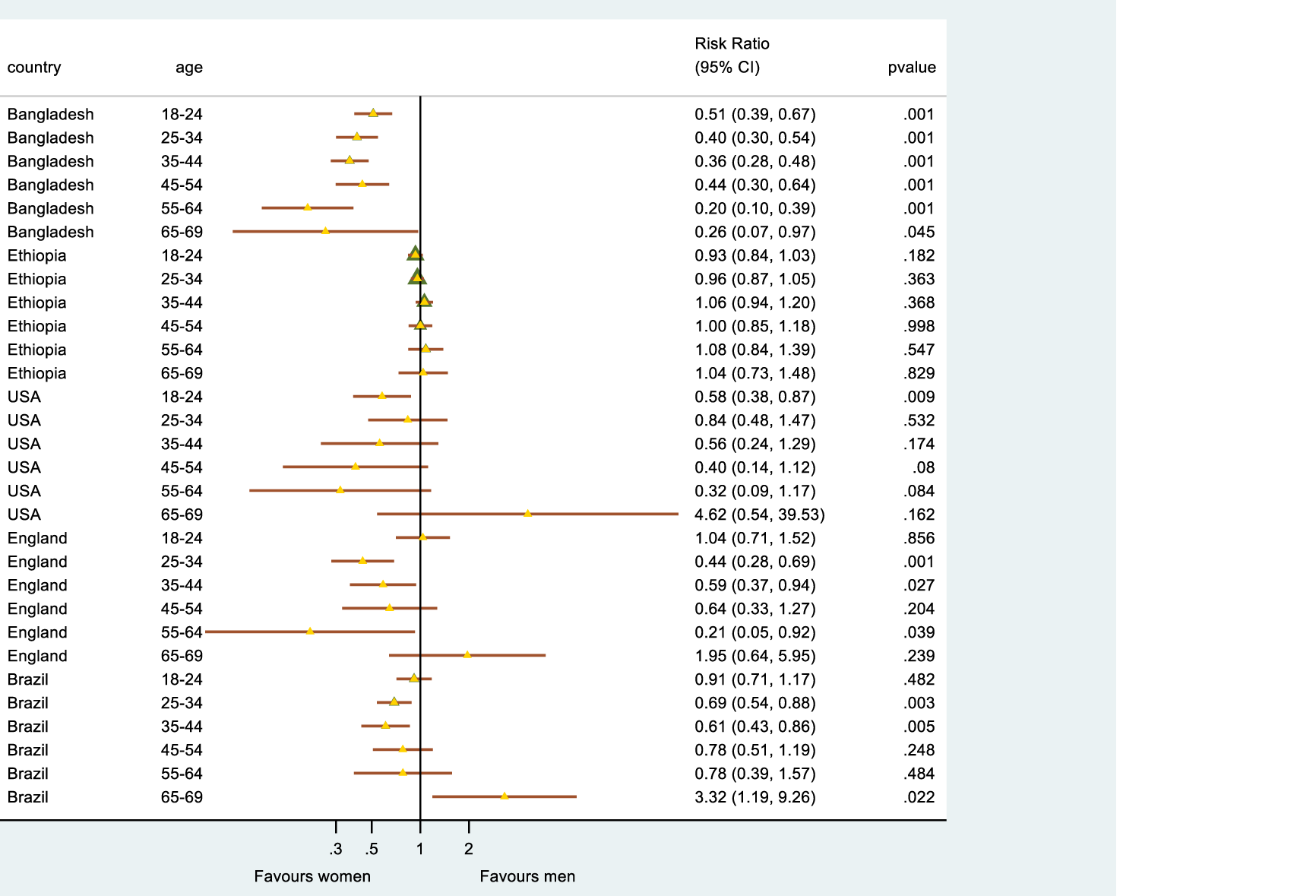
Sex specific ideal CVH risk by age group for each country - Based on five metrics (BP, BMI, cholesterol, glycaemia, smoking)

## Discussion

This research sought to harmonize, analyze, and compare CVH data from nationally representative adult surveys of five diverse high- to low-income countries to test the hypothesis that the decline in CVH with age is universal. While comparable data were lacking for diet and sleep, analysis of the remaining CVH metrics (BP, glycaemia, BMI, smoking, cholesterol, and PA) supported our hypothesis and showed that the prevalence of high CVH scores declined across the life course in all five countries. Further, our hypothesis that the starting point and the tempo of decline may differ between high-, middle- and low-income countries was also supported within this analysis.

Our findings in Brazil that 56.2% of adults had high CVH scores using five CVH metrics are similar to those previously published from the Brazilian Health Survey (PNS 2013). Dias Moreira et al. (2020) reported that among 4,585 Brazilian adults from the PNS survey (mean age 43.2 years ± 23.7 SD), 56.7% had ideal CVH scores based on four or more LS7 CVH metrics in the ideal range.^14^ The results from Brazil were similar to those in the US NHANES sample (56.1% of adults scored high for CVH or ≥8/10 based on the five CVH metrics). While our analysis used the 2017-2020 pre-pandemic NHANES dataset, analysis using earlier NHANES data suggests a much lower prevalence of ideal CVH (17-18% of adults ≥20y with ≥5 of the LS7 metrics at ideal levels; 5% with ≥6 ideal CVH metrics; and 2% or less with all of the LS7 CVH metrics at ideal levels).^4, 32, 33^ However, this may reflect differences in scoring, for example, between counts of the number of individual metrics classified as ideal which typically generates lower prevalence estimates of good or high CVH when compared to the allocation of scores to each metric which are summed to give an overall high, moderate, or low CVH rating. More recent analysis in NHANES data employing this latter approach with all of the LE8 CVH metrics produces more similar estimates with a mean score of 65.2/100 in US adults ≥20yrs,^34^ compared to our observed mean CVH scores of 6.4/10 or 7.8/12. Additionally, as we have shown, the population prevalence of a high CVH score increases as the number of metrics used decreases, such that with six metrics we found 40.4% of US adults scored high (≥9/12), while with five metrics this increased to 56.1% of US adults with high CVH scores (≥8/10). This is similar to data from a larger US adult sample showing 43.5% high CVH score when using six metrics (≥5/6, n=177,421).^35^ Among the US sample, we did observe that the BMI metric scored particularly low with 74% of adults scored as moderate or low (overweight or obese). This is consistent with previous NHANES analysis showing 56% overweight/obese in 1998-94, increasing to 68% in 2005-10.^4^

Previous analysis conducted in Bangladesh, also using the 2018 STEPS survey, showed 43% of adults had at least 5 of the LS7 metrics at ideal levels,^36^ corresponding to twice the level reported in US adults (17-18%) using the same approach.^32, 33^ While we did not find quite this magnitude of difference, possibly as diet was not evaluated, we did observe Bangladesh scored higher than the US on total CVH score using either six or five metrics, and on each CVH metric individually with the exception of smoking, where Bangladesh had the highest proportion of current smokers (24%). In contrast, England had the lowest prevalence of high CVH scores for smoking, primarily due to a higher prevalence of former smokers (33%). This HSE data was collected in 2016, post implementation of stringent UK tobacco control policies and may reflect the success of these.^37^ However, our finding that 40% of UK adults in 2016 achieved a high CVH score based on six metrics, is much higher than the 14% found earlier in over 300,000 adults from the UK Biobank study (2006-2010) using a similar scoring method and all of the LS7 metrics.^38^ This difference may be due to both the inclusion of diet, for which fewer adults score well, and the Biobank sample being older (37-73 years).

We were unable to find evidence of the previous application of the CVH score in Ethiopia for comparison. Our prevalence of overweight and obesity (6.5%) in Ethiopia is lower than many previous findings which suggest a pooled prevalence of overweight and obesity at approximately 24%.^39^ This difference may in part be due to sampling strategy (national sampling vs urban samples alone which tend to show higher BMI) and sample size. This may also be influenced by the high number of underweight individuals in our study that were classified as high CVH scores for BMI due to the CVH scoring including all individuals with BMI<25 kg/m^2^. While we did evaluate the impact of this on the total CVH score and found it to be negligible overall, application of a lower threshold (BMI 18.5 kg/m^2^, based in the WHO classification for underweight) for the high BMI CVH category may be needed, especially as being underweight may confer additional risk for CVD.^40^ Furthermore, when applying scores for BMI in populations in other geographical locations or with different ethnicities, the BMI cut-offs may need to be revised as the level that is predictive of cardio-metabolic risk may be lower.^41^ As such it is possible that our analysis in Bangladesh especially may present a more optimistic situation than is actually the case though the lower levels of obesity (defined as BMI<25 kg/m^2^) in Ethiopia and Bangladesh also presented with lower levels of related hyperglycaemia and hypercholesterolemia, potentially explaining the apparent higher prevalence of high CVH in these lower-income countries. It is important to compare other health behaviors across countries, as physical activity also varied widely and, with the exception of Brazil with a different PA data collection method, generally decreased as country income level increased. This may reflect the reductions in energy expenditure associated with occupation, transport and domestic activities observed in higher income countries over the past decades that are not (yet) observed to the same extent in lower income countries.^42^

We also found that just 5% of adults in Ethiopia use tobacco compared to 13-17% in higher income countries and 24% in Bangladesh. This may explain in part why the percentage of deaths from non-communicable disease (NCD), including CVD, may follow the overall CVH scoring pattern and decline as country income level declines (88% in the UK and US, 75% in Brazil, 70% in Bangladesh and 43% in Ethiopia^25^), while Bangladesh NCD mortality rates are more similar to the higher income countries than to Ethiopia. Additionally, the percentage of deaths from maternal and child health, nutrition or communicable disease remains high at 45% in Ethiopia compared to 5% in the US.^25^ In addition to differing causes of mortality across the countries, differences were observed in access to health screening and treatment, with lower rates of detection and treatment for hypertension in lower income compared to higher income countries. These findings reinforce previous calls for health systems strengthening in LMICs to prevent a range of chronic diseases.^43^

Furthermore, it is possible that other variables not yet captured in the CVH scoring may influence relative NCD mortality and morbidity rates. For example, UK adults consume 11.5 liters of alcohol per capita while in Bangladesh this is below 0.5 liters per capita,^44^ while evidence is growing on the impact of both positive and negative psychosocial factors on cardiovascular health and disease (including optimism, mindfulness, resilience, and hope as well as depression, perceived stress at home or work, low locus of control, and major life events) across low- to high-income countries.^45, 46^ In addition, it should be acknowledged that among children and adults living in low-income countries, rheumatic heart disease (RHD) is a common cause of acquired heart disease.^47, 48^ As such, the LS7/LE8 metrics, which focus on risk factors for atherosclerotic CVD may not fully capture true CVH in these settings.

The results of this secondary data analysis should be viewed within the limitations of this study. For example, surveys were not all conducted at the same time with the year of data collection ranging from 2013 (Brazil) up to pre-pandemic 2020 (US). The lack of availability of both sleep and dietary data is a further limitation in this analysis. Challenges harmonizing dietary data collected from such different countries may have been expected as a previous secondary data analysis showed this could not be achieved for population surveys from six countries within Europe.^49^ While the dietary metrics included in the initial LS7 CVH scoring were fairly simple,^2^ these data are not routinely collected across populations in a way permitting harmonization. To counteract such challenges, the revised LE8 CVH scoring guidelines proposed a new method for assessing dietary quality that could be employed for both rapid individual assessment in clinical settings and for population-level surveys.^3^ However, this has yet to be widely deployed, especially in LMICs. Sleep could also not be harmonized as not all countries included sleep assessment. Future surveys could include this measure as the single question proposed by LE8: “On average, how many hours of sleep do you get per night?”. The validity of simple sleep questions in comparison to objectively measured sleep data has proved challenging in the past^50, 51^ and may need further research, especially across different regions, languages, and cultures. A further limitation lies in the physical activity analysis, with Brazil using PAVS, a PA questionnaire that differed from other country approaches. The lower levels of PA reported using PAVS compared to instruments that explore multiple activity domains or types has been reported previously, where PAVS was found on average to record 86 minutes less MVPA per week.^52^ Brazil reported lower MVPA in our study; possibly Brazil does have lower prevalence of achieving the PA recommendations. For example, a national study in Brazil using a PA instrument that queried multiple activity domains found that in 2013, 50% of the population met the PA guidelines^53^ and would therefore have met the criteria for high CVH PA categorization. While this is higher than the 17% high CVH PA categorization we observed, it remains lower than the other four countries included in this analysis.

The wide variation in missing data across the surveys is another limitation, with NHANES presenting the largest amount of missing data. An analysis comparing the NHANES analytical sample with eligible NHANES participants who were excluded due to missing data showed that there were no significant differences in the proportions of men and women, or in levels of education but there was a significant difference in age between these groups with excluded participants being younger (mean age in analytical sample 46y ± 15.0 and in the excluded sample 42y ± 15.1 (p<0.001)). This may be due, in part, to how cholesterol medication is recorded in this survey. For example, to understand treatment use, all countries ask questions that generally follow the care cascade (have you been measured, diagnosed, prescribed treatment and are you taking treatment?). However, in the US, primary and secondary prevention of CVD does not require high cholesterol levels to indicate statins with statins being used according to CV risk and age, not only cholesterol levels.^54^ Indeed, this was observed in our data where the US showed the second highest mean total cholesterol level among the five countries but the highest percentage on treatment for lipids (more than twice that of England).

The significant differences between countries in population age structure, although representative of national demographics, also present a challenge for such comparative analysis. For example, the age-specific trajectories of good (high) CVH across the life course were lower at all ages for Brazil whether this was based on six or five of the CVH metrics indicating a relatively lower prevalence of good CVH across the adult age range when compared to the other countries. However, the mean CVH score in Brazil (when PA is excluded) appears higher than the scores from England and the US, possibly due to the younger age distribution in PNS compared to NHANES and HSE. The scoring of each CVH metric (presented in Table 3), though not age-adjusted, was weighted to each survey in order to present and compare the population prevalence of poor CVH related to each CVH metric between countries. However, the patterns observed in the data tables and in the figures appear to agree that CVH assessed in these five countries and using these methods declines in populations as country income level increases. Further analysis is needed across a greater number of countries to confirm this.

Furthermore, the lack of consistent information available for the CVH metrics sometimes constrained the analysis that could be employed. For example, the WHO-STEPS surveys collected information on the use of traditional medicines for BP, cholesterol, or glucose; however, this was not included in the analysis as not all surveys had these data and because the effects of these medications may vary and could not be predicted. Also, surveys had information on current and former smoking, but few had data on broader tobacco exposure or use of other tobacco products, data required under LE8. These newer guidelines are different in some important respects. For example, the terms ‘ideal’, ’intermediate’, and ’poor’ were set aside and replaced by a score from 0-100 for each of the eight metrics and overall CVH score, such that 80% or more of the composite score is classified as high CVH.

A strength of this analysis is the comparison across a highly diverse group of 5 low- to high-income countries including a total combined analytical sample of over 28,000 adults, with selected countries at varying stages of urban, economic, and nutritional transition. While over 80% of people in Brazil, US and England live in urban areas, in Bangladesh and Ethiopia, urban dwellers still form less than one quarter of the population.^55^ Despite increasing urbanization, there has been relatively little change in the food supply in Ethiopia over the last 50 years.^56^

A further strength in this analysis is the general agreement between our findings and those of the Global Burden of Disease Study (GBD, 2019).^57^ Regarding the high apparent CVH in Ethiopia, data from country profiles provided by the GBDS are consistent with our findings. In Ethiopia, only 4 of the 10 leading contributors to death and disability were CVH metrics, and they ranked low in importance (BMI being lowest of 10), while in Brazil, the US, and the UK, CVH metrics comprised the top 5 contributors with a 6th at lower levels; the picture for Bangladesh was intermediate.

Despite this apparent alignment, the utility and potential adaptation of CVH scoring for use across multiple countries requires further exploration. Future research utilizing population surveys from multiple countries and regions is needed. To facilitate this, the formulation of a single protocol for data collection is needed that considers the contextual challenges of low resource settings. A single questionnaire that includes all relevant questions for CVH assessment, with methods that can be applied across high- and low-income settings will allow validations of this approach in different countries and regions as well as support global comparisons. Harmonized collection of data on dietary quality across countries is likely to require special consideration.

While there are important differences between the included countries, our analysis suggests that the decline in high CVH seen from adolescence across the adult life course may be universal. Differences observed between countries and occurring already at age 18 years may indicate that strategies to maintain higher CVH in this young adult age range may be more successful sooner in lower than in higher income countries, where CVH is already lower at this age. This indicates the need for earlier life interventions in HICs, with a major focus on attaining and maintaining a healthy body weight from childhood across the life course. Our data on the individual CVH metrics across the countries may be useful to inform other country specific intervention strategies. For example, efforts in Bangladesh to reduce tobacco use could utilize learnings from effective tobacco control policies observed in the UK. While in Ethiopia, policies envisioned by Strasser, Popkin and others^58, 59^ aimed at protecting the whole of society as transitions occur to support the primordial prevention of CV risk are likely to be beneficial. Although approximately 80% of global CVD related deaths occur in LMICs, based on our analysis the age-specific CVH rates are higher in LMICs compared to HICs. Access to and quality of care may partly explain this mortality disparity, however future studies need to assess the association of CVH with cause-specific mortality in these populations.

## Conclusions

Our harmonization of data on CVH metrics from nationally representative population surveys in 5 widely diverse low- to high-income countries reveals a consistent pattern of decline in overall CVH with age, supporting our hypothesis that this may be universal across countries. Differences in prevalence of high CVH already at age 18 suggests important differences in starting levels from birth, tempos of decline, and inflection points to be considered in CVH promotion strategies in each country. Lack of dietary data suitable for harmonization remains a critical problem to be resolved as new standardized methods for population studies of CVH are designed. Given the rapid urban, economic and nutrition transitions taking place in LMICs, targeted and contextually relevant interventions to change CVH trajectories are needed now to increase prevalence of high CVH at all ages, globally.

## Data Availability

All data produced in the present study are available upon reasonable request to the authors.

## Acknowledgments

This paper uses data from the Bangladesh 2018 and Ethiopia 2015 STEPS surveys, implemented by the Bangladesh National Institute of Preventive and Social Medicine (NIPSOM) and the Ethiopian Public Health Institute (EPHI) respectively with the support of the World Health Organization. Kavita Singh is supported by the Fogarty International Centre, National Institutes of Health (NIH), United States (grant award: 1K43TW011164).

## Sources of Funding

This work was conducted with funding support from the Institute for Global Health, Northwestern University [Catalyzer Award No. 1005]; from the DSI-NRF Centre of Excellence in Human Development hosted at the University of the Witwatersrand in South Africa, and the support of the University of the Witwatersrand research office.

## Disclosures

None to declare.

## Notes

### Competing Interest Statement

The authors have declared no competing interest.

### Funding Statement

This study was funded with support from the Institute for Global Health, Northwestern University [Catalyzer Award No. 1005]; from the DSI-NRF Centre of Excellence in Human Development hosted at the University of the Witwatersrand in South Africa, and the support of the University of the Witwatersrand research office.

### Author Declarations

Ethics committee/IRB of the University of the Witwatersrand Medical Human Research Ethics Committee gave ethical approval for this work. [Ref: M220437]

