## Supplementary material for "Comparison of cardiovascular health profiles across population surveys from five high- to low-income countries": Suppl Figure 1 and Suppl Table 1

**Supplementary Figure 1:** Current medication use for high blood pressure (BP), cholesterol, and diabetes was extracted from care cascade data available for each country - demonstrated here for BP.

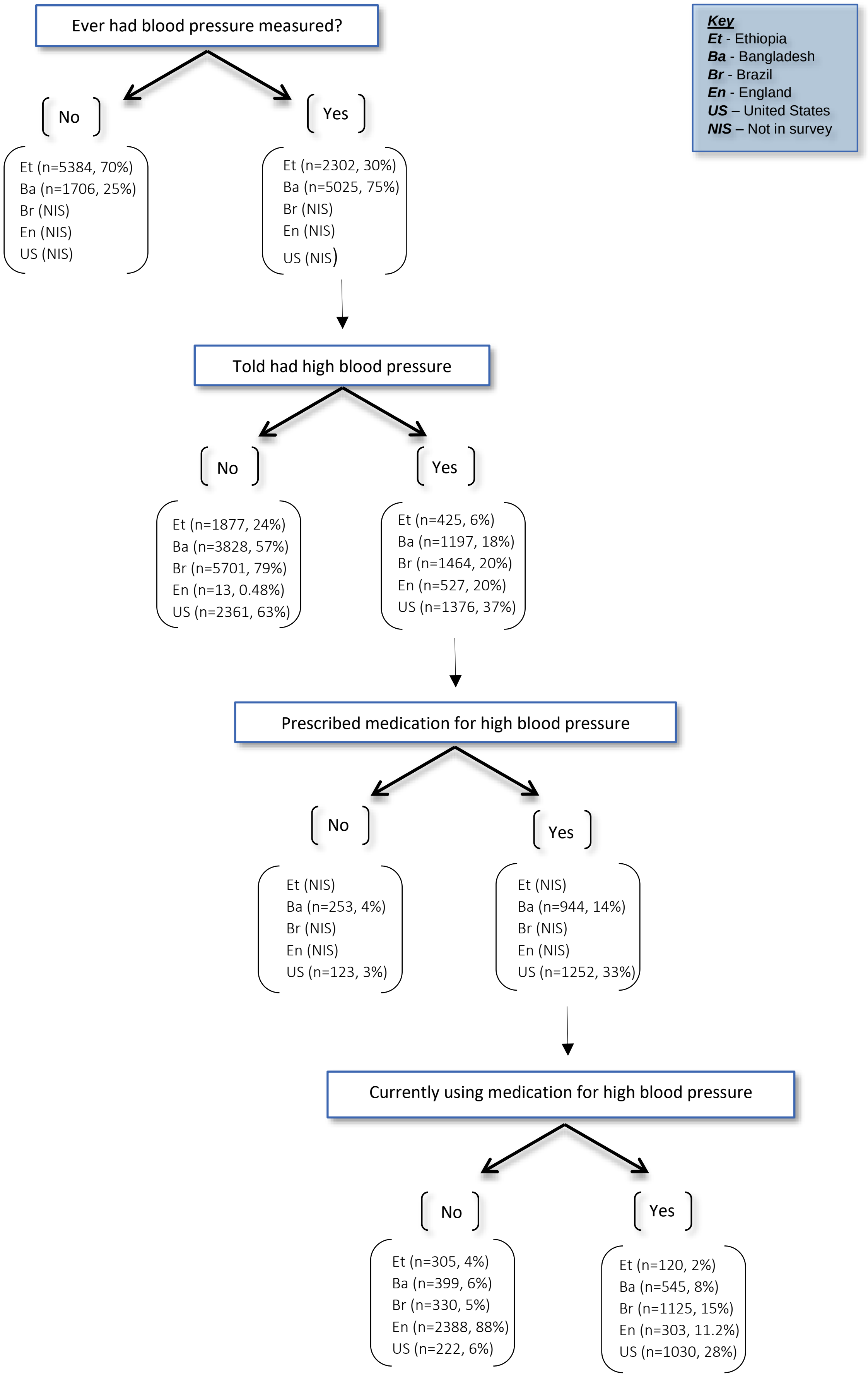

**Supplementary Table 1:** Harmonization of cardiovascular health metrics and other key variables. (Adapted from Schaap et al.). The recoding of variables was conducted within each country dataset with the recoded categories then combined in a multi-country dataset for further analysis. Syntax shown is using IBM® SPSS® Statistics, Version 28 (IBM Corporation, Armonk, New York).

**CARDIOVASCULAR HEALTH METRICS**

| COUNTRY | QUESTION (variable name) | VALUE | CODE ASSIGNED |
|---|---|---|---|
| | **Country** | (country) | 1. Bangladesh; 2. Ethiopia; 3. US; 4. England; 5. Brazil |
| | **Cardiovascular health score** | (cvh_score); (cvh_cat) | 0. Low; 1. Intermediate; 2. High |

***Body mass index:*** Weight (kg) and height (m) were measured in all five surveys, with BMI calculated as weight in kg divided by height in meters squared already in the survey dataset (NHANES, HSE) or calculated from the survey height and weight variables (WHO-STEPS, PNS). Both NHANES and WHO-STEPS have measurement protocols specifying participants should remove shoes, headwear (when possible) and heavy items from pockets or outdoor clothing, based on WHO standard guidelines.[1] Trained data collection staff in Bangladesh, NHANES and HSE used portable stadiometers and digital weighing scales to measure height and weight respectively.[2 3] In Ethiopia, an electronic Growth Management Scale was used to measure both height and weight.[4] The PNS survey conducted two measurements of height and weight, using the average of the two when values were equal or differed by ≤1 cm for height and ≤0.5 kg for weight.[5] All surveys used measurement techniques based on the WHO recommended techniques for anthropometry.[1]

| | **Body mass index** | **(bmi_cat)** | 0. ≥30 $kg/m^2$; 1. 25-29.9 $kg/m^2$; 2. <25 $kg/m^2$ |
|---|---|---|---|
| **Bangladesh** | Weight (kg) (m12) and height (cm) (m11) measured | height=m11/100 bmi=m12/$(height)^2$ | Recoded as categorical variable:<br>IF (bmi>=30) bmi_cat=0.<br>IF (bmi>=25 AND bmi<30) bmi_cat=1.<br>IF (bmi<25) bmi_cat=2. |
| **Ethiopia** | Weight (kg) (m12) and height (cm) (m11) measured | height=m11/100 bmi=m12/$(height)^2$ | |
| **US** | BMI calculated (BMXBMI) | bmi=(BMXBMI) | |
| **England** | BMI calculated (BMI) | bmi=(BMI) | |
| **Brazil** | Weight (kg) (Z004) and height (cm) (Z005) measured | height=Z005/100 bmi=Z004/$(height)^2$ | |

***Blood pressure:*** To assess BP, both systolic and diastolic BP measurements and current antihypertensive medication use are required. All surveys recorded current use of medication for hypertension and used an automated BP monitor to measure brachial BP three times, with protocols closely aligned to the International Society of Hypertension measurement guidelines (participant seated, with legs uncrossed and upper arm rested at the level of the heart with a rest of at least 3 minutes before the measurement and of at least one minute between measures).[6] Bangladesh used a BP–BOSO–Medicus Control with universal cuff, taking measurements from the left arm.[7] Ethiopia used the same device with measurements taken on the right arm.[4] NHANES used an Omron HEM–907XL device with BP measurements taken on the right arm unless this was not possible.[8] England's HSE used an Omron automated device on the right arm,[9] while Brazil used a calibrated digital device on the left arm.[10] Guidelines recommend discarding the first reading and averaging the second and third for analysis.[6] However, the Brazil dataset included average SBP and DBP from all three readings, so the same approach was applied for all country data.

| | **Blood pressure** | **(mean_bp_cat)** | 0. SBP ≥140 or DBP ≥90 mmHg (treated/untreated); 1. SBP 120-139 or DBP 80-89 mmHg (untreated); 1. SBP <140 or DBP <90 mmHg (treated); 2. SBP <120 or DBP <80 mmHg (untreated) |
|---|---|---|---|
| **Bangladesh** | Systolic (m4a, m5a, m6a) and diastolic BP (m4b, m5b, m6b) measured on the **left arm** (mmHg)<br>1. Was your blood pressure measured? (h1)<br>2. Were you told you have/had high blood pressure? (h2a)<br>3. Have you ever taken drugs/medications for raised blood pressure prescribed by a doctor/health worker? (hx1)<br>4. In the past two weeks, have you taken any drugs (medication) for raised blood pressure prescribed by a doctor or other health worker? (h3) | mean_sbp=(m4a+m5a+m6a)/3<br>mean_dbp=(m4b+m5b+m6b)/3<br>Unless otherwise stated, all questions were answered as: 1. Yes; 2. No | curr_tx_bp = Current medication usage for high blood pressure (1=YES, 2=NO; **Suppl. Figure 1**)<br>Recoded as categorical variable:<br>IF (mean_sbp>=140) OR (mean_dbp>=90) HTN_cat=0.<br>IF (mean_sbp<120) AND (mean_dbp<80) HTN_cat=2.<br>RECODE HTN_cat (0=0) (2=2) (MISSING=1) (ELSE=1).<br>IF (HTN_cat=0) mean_bp_cat=0.<br>IF (curr_tx_bp=1) AND (HTN_cat>0) mean_bp_cat=1.<br>IF (curr_tx_bp=2) AND (HTN_cat=1) mean_bp_cat=1.<br>IF (curr_tx_bp=2) AND (HTN_cat=2) mean_bp_cat=2. |
| **Ethiopia** | Systolic (m4a, m5a, m6a) and diastolic BP (m4b, m5b, m6b) measured on the **left arm** (mmHg)<br>1. Was your blood pressure measured? (h1)<br>2. Were you told you have/had high blood pressure? (h2a)<br>3. In the past two weeks, have you taken any drugs (medication) for raised blood pressure prescribed by a doctor or other health worker? (h3) | mean_sbp=(m4a+m5a+m6a)/3<br>mean_dbp=(m4b+m5b+m6b)/3<br>Unless otherwise stated, all questions were answered as: 1. Yes; 2. No | |
| **US** | Systolic (BPXOSY1, BPXOSY2, BPXOSY3) and diastolic BP (BPXODI1, BPXODI2, BPXODI3) measured on the **right arm** (mmHg)<br>1. Were you ever told you have high blood pressure? (BPQ020)<br>2. Are you taking any prescription medication for hypertension? (BPQ040A) | mean_sbp=(BPXOSY1+BPXOSY2+BPXOSY3)/3<br>mean_dbp=(BPXODI1+BPXODI2+BPXODI3)/3<br>Unless otherwise stated, all questions were answered as: 1. Yes; 2. No | |

| | | |
|---|---|---|
| | 3. Are you currently taking medication for high blood pressure? (BPQ050A) | |
| **England** | Systolic (SYS1OM, SYS2OM, SYS3OM) and diastolic BP (DIAS1OM, DIAS2OM, DIAS3OM) measured on the **right arm** (mmHg)<br>1. Have you ever had high blood pressure? (EverBP)<br>2. Were told by a doctor/nurse you had high blood pressure? (DocBP)<br>3. Are you taking any drugs for blood pressure? (bpmedd2) | mean_sbp=(SYS1OM+SYS2OM+SYS3OM)/3<br>mean_dbp=(DIAS1OM+DIAS2OM+DIAS3OM)/3<br>Unless otherwise stated, all questions were answered as: 1. Yes; 2. No<br>Q3 0. Not taking drug; 1. Taking drug |
| **Brazil** | Average systolic (W00407) and average diastolic BP (W00408) from 3 readings on the **left arm** (mmHg)<br>1. Were you ever diagnosed with high blood pressure? (Q002)<br>2. In the last two weeks, have you taken medication for high blood pressure? (Q006) | mean_sbp=W00407 mean_dbp=W00408<br>Q1 1. Yes; 2. Only during pregnancy; 3. No<br>Q2 1. Yes; 2. No |

***Cigarette smoking:*** To assess tobacco use, data is required on current or previous smoking behaviour, with the recent update including use of vaping or e-cigarettes and second-hand tobacco exposure through living with an active smoker in the home.[11] All surveys collected data to determine self-reported current or previous smoking behavior. Bangladesh included data on daily or ever use of electronic cigarettes but had no information on exposure to tobacco smoke within the home. Brazil and Ethiopia asked about exposure to cigarette smoke within the home but had no information on vaping or e-cigarette use. While NHANES had data on e-cigarette use and exposure to second-hand smoke in multiple locations outside of the home, there was no data on whether the participant lived with an active smoker inside their home. Only the HSE had sufficient data to assess the full range of indicators recommended by AHA for tobacco use or exposure. As such, the decision was taken to harmonize tobacco use based on the 2010 categorization only of current or previous smoking behavior.[12]

| | **Smoking** | **(smoking)** | 0.Yes; 1. Former ≤12 months; 2. Never or quit >12 months |
|---|---|---|---|
| **Bangladesh** | Self-reported – TQS<br>1. Do you currently smoke any tobacco products such as cigarettes, bidis, hookah, cigars, or pipes? (t1)<br>2. In the past, did you ever smoke any tobacco products? (t8)<br>3. In the past, did you smoke daily tobacco products? (t9)<br>4. How long ago did you stop smoking? – *Data not available* | Unless otherwise stated, all questions were answered as: 1. Yes; 2. No | Recoded as:<br>IF (t1=1 AND t2=1 AND t8=1 AND t9=1) smoking=0.<br>IF (t8=1 AND t9=1) smoking=1.<br>IF (t1=2 AND t2=2 AND t8=2 AND t9=2) smoking=2. |
| **Ethiopia** | Self-reported – TQS | Unless otherwise stated, all questions were answered as: 1. Yes; 2. No | |

| | | | |
|---|---|---|---|
| | 1. Do you currently smoke any tobacco products such as cigarettes, bidis, hookah, cigars, or pipes? (t1)<br>2. In the past, did you ever smoke any tobacco products? (t8)<br>3. In the past, did you smoke daily tobacco products? (t9)<br>4. How long ago did you stop smoking? – *Data not available* | | |
| **US** | Self-reported – ACASI<br>1. Have you smoked at least 100 cigarettes in your entire life? (SMQ020)<br>2. Do you now smoke cigarettes…? (SMQ040)<br>3. How long has it been since you quit smoking cigarettes? – *Data not available* | Q1 1.Yes; 2.No<br>Q2 1. Every day; 2. Some days; 3.Not at all | Recoded as:<br>IF (SMQ020 = 1) smoking=0.<br>IF (SMQ020 = 2) smoking=2.<br>IF (SMQ040 = 1) smoking=0.<br>IF (SMQ040 = 2) smoking=1.<br>IF (SMQ040 = 3) smoking=2. |
| **England** | Self-reported<br>1. Have you ever smoked a cigarette, a cigar, or a pipe? (smkevr)<br>2. Do you smoke cigarettes at all nowadays? (cignow)<br>3. Have you ever smoked cigarettes? (cigevr)<br>4. How long ago did you stop smoking? – *Not able to harmonize* | Unless otherwise stated, all questions were answered as: 1. Yes; 2. No | Recoded as:<br>IF (smkevr=1 OR cignow=1 OR cigevr=1) smoking=0.<br>IF (smkevr=1 AND cignow=2 AND cigevr=1) smoking=1.<br>IF (smkevr=2 AND cignow=2 OR cigevr=2) smoking=2. |
| **Brazil** | Self-reported<br>1. Do you currently smoke any tobacco products? (P050) | 1. Yes, daily.<br>2. Yes, less than daily.<br>3. No | Recoded as:<br>IF (P050=1 OR P050=2) smoking=0.<br>IF (P050=2 OR P050=3) smoking=1.<br>IF (P050=3) smoking=2. |

***Physical activity:*** To assess this metric, the minutes of moderate- and vigorous-intensity PA (MVPA) in a typical week are required, with the updated scoring suggesting use of the PAQ-K questionnaire[11] used within the NHANES survey. This questionnaire is similar to both the General Practice Physical Activity Questionnaire (GPAQ;[13] used in Bangladesh and Ethiopia) and the International Physical Activity Questionnaire (IPAQ;[14] used in England). Each of these instruments examines the frequency and duration of MVPA across the domains of work, active travel and leisure time allowing the calculation of the required weekly MVPA. In contrast, Brazil used a shorter questionnaire similar to the Physical Activity Vital Sign (PAVS[15]), that asked about the duration and frequency of all types of physical exercise or sport. As this allowed the calculation of total PA per week, which in theory would include MVPA, it was decided to harmonize this variable across the surveys.

| | | | |
|---|---|---|---|
| | **Physical activity** | **(phys_act)** | 0.None; 1. 1-149 min/week moderate intensity or 1-74 min-week vigorous intensity or 1-149 moderate + vigorous; 2. ≥150 min/week |

| | | | |
|---|---|---|---|
| | | | moderate intensity or ≥75 min/week vigorous intensity or ≥150 min/week moderate + vigorous |
| **Bangladesh** | Self-reported – GPAQ<br>Vigorous activity at work (p1 – yes/no)<br>• Days per week (p2); Hours per day (p3a); Minutes per day (p3b)<br>Moderate activity at work (p4 – yes/no)<br>• Days per week (p5); Hours per day (p6a); Minutes per day (p6b)<br>Active transport (p7 – yes/no)<br>• Days per week (p8); Hours per day (p9a); Minutes per day (p9b)<br>Vigorous leisure (p10 – yes/no)<br>• Days per week (p11); Hours per day (p12a); Minutes per day (p12b)<br>Moderate leisure activity (p13 – yes/no)<br>• Days per week (p14); Hours per day (p15a); Minutes per day (p15b)<br>Sedentary time<br>• Hours per day (p16a); Minutes per day (p16b) | | Physical activity was coded according to the standardized WHO GPAQ cleaning code and categorized as:<br><br>**Poor** – Physical inactive<br>**Intermediate** – 1-149 min/week moderate intensity or 1-74 min-week vigorous intensity or 1-149 moderate + vigorous.<br>**Ideal** – ≥150 min/week moderate intensity or ≥75 min/week vigorous intensity or ≥150 min/week moderate + vigorous |
| **Ethiopia** | Self-reported – GPAQ<br>Vigorous activity at work (p1 – yes/no)<br>• Days per week (p2); Hours per day (p3a); Minutes per day (p3b)<br>Moderate activity at work (p4 – yes/no)<br>• Days per week (p5); Hours per day (p6a); Minutes per day (p6b)<br>Active transport (p7 – yes/no)<br>• Days per week (p8); Hours per day (p9a); Minutes per day (p9b)<br>Vigorous leisure (p10 – yes/no)<br>• Days per week (p11); Hours per day (p12a); Minutes per day (p12b)<br>Moderate leisure activity (p13 – yes/no)<br>• Days per week (p14); Hours per day (p15a); Minutes per day (p15b)<br>Sedentary time<br>• Hours per day (p16a); Minutes per day (p16b) | | |
| **US** | Self-reported – PAQ (based on GPAQ)<br>Vigorous work activity (PAQ605 – yes/no) | | |

| | | | |
|---|---|---|---|
| | • Number of days (PAQ610); Minutes vigorous-intensity work (PAD615)<br>Moderate work activity (PAQ620)<br>• Number of days (PAQ625); Minutes moderate-intensity work (PAD630)<br>Walk or bicycle (PAQ635)<br>• Number of days (PAQ640); Minutes walk/bicycle for transportation (PAD645)<br>Vigorous recreational activities (PAQ650)<br>• Number of days (PAQ655); Minutes vigorous recreational activities (PAD660)<br>Moderate recreational activities (PAQ665)<br>• Number of days (PAQ670); Minutes moderate recreational activities (PAD675)<br>Sedentary time<br>• Minutes sedentary activity (PAD680) | | |
| **England** | Self-reported – IPAQ<br>Combined variable for moderate-vigorous physical activity (MVPA) (recs12_2) | 1. **Inactive:** Less than 30 min/week of MPA, less than 15 min/week of VPA, or a combination<br>2. **Low or some physical activity:** 30-59 (low) or 60-149 (some) min/week of MPA, 15-29 (low) or 30-74 (some) min/week of VPA, or a combination<br>3. **Meets MVPA guidelines:** 150 min/week of MPA, or 75 min/week of VPA, or a combination | Recoded as:<br>IF (recs12_2=1) phys_act=0.<br>IF (recs12_2=2) phys_act=1.<br>IF (recs12_2=3) phys_act=2. |
| **Brazil** | Self-reported<br>1. How many days per week do you usually practice any physical activity/sport? (P035)<br>2. On these days that you practice physical activity/sport, how long does it last? (minutes) (P03702) | Total minutes of activity (total_minutes) = P035 x P03702 | Recoded as:<br>IF (total_minutes <= 0) phys_act=0.<br>IF (total_minutes >= 1 AND total_minutes <= 149) phys_act=1.<br>IF (total_minutes >= 150) phys_act=2. |

*Cholesterol:* To assess CVH, the LS7 CVH scoring requires a measure of total cholesterol and of treatment use for raised cholesterol.[12] In the LE8 updated CVH scoring, a measurement of HDL-cholesterol is also required in addition to total cholesterol to calculate non-HDL cholesterol, specifying the use of either fasting or non-fasting plasma.[11]

Bangladesh measured total and HDL cholesterol in fasting venous plasma samples using an autoanalyser (Elitech® Selectrao Pro M). Ethiopia measured total and HDL cholesterol using a CardioCheck PA Analyser with fasting capillary blood samples. NHANES measured total and HDL cholesterol in fasting serum samples (Roche/Hitachi Cobas 6000 Chemistry Analyser), while HSE measured total and HDL cholesterol in non-fasting serum samples, using the Cholesterol Oxidase assay method on a Roche Cobas 702 analyser. Brazil also measured total and HDL cholesterol in non-fasting serum samples using an automated enzymatic/colorimetric method. Current self-reported medication use for raised cholesterol was assessed in all surveys. As there may be greater variability between capillary and venous samples for HDL cholesterol than for total cholesterol,[16] the decision was taken to harmonize total cholesterol values only.

| | **Total cholesterol** | **(total_chol)** | 0. ≥240 mg/dl (treated/untreated); 1. <240 mg/dl (treated); 1. 200-239 mg/dl (untreated); 2. <200 mg/dl (untreated) |
|---|---|---|---|
| **Bangladesh** | Measured in fasting plasma (mg/dL) (b8)<br>1. Was your cholesterol measured? (h12)<br>2. Were you told you have/had high cholesterol? (h13a)<br>3. Have you taken meds for high cholesterol prescribed by a doctor/health worker? (hx9)<br>4. Have you taken meds for high cholesterol in the past 2 weeks? (h14) | b8=total_chol_measure<br>Unless otherwise stated, all questions were answered as: 1. Yes; 2. No | curr_tx_chol=Current medication usage for high cholesterol (1=YES, 2=NO; **Suppl. Figure 1**)<br><br>Recoded as categorical variable:<br>IF (total_chol_measure>239) total_chol=0.<br>DO IF (curr_tx_chol=1).<br>IF (total_chol_measure<240) total_chol=1.<br>END IF.<br>DO IF (curr_tx_chol=2).<br>IF (total_chol_measure >199) AND (total_chol_measure <240) total_chol=1.<br>IF (total_chol_measure <200) total_chol=2.<br>END IF. |
| **Ethiopia** | Measured in fasting plasma (mg/dL) (b8)<br>1. Was your cholesterol measured? (h12)<br>2. Were you told you have/had high cholesterol? (h13a)<br>3. Have you taken meds for high cholesterol in the past 2 weeks? (h14) | | |
| **US** | Measured in fasting serum (mg/dL) (LBXTC)<br>1. Ever had your cholesterol checked? (BPQ060)<br>2. Were you told you have/had high cholesterol? (BPQ080)<br>3. Are you taking any prescription medication for high cholesterol? (BPQ090)<br>4. Are you currently taking prescribed medicine for high cholesterol? (BPQ100D) | LBXTC=total_chol_measure<br>Unless otherwise stated, all questions were answered as: 1. Yes; 2. No | |
| **England** | Measured in non-fasting serum (mmol/L) (Cholest)<br>1. Are you currently taking any lipid lowering prescribed medication? (lipid2)<br>2. Any prescribed lipid-lowering medications taken in the last 7 days? (LipidTakg2) | chol_mg_dl= Cholest * 38.67<br>chol_mg_dl=total_chol_measure<br>Q1 0. Not taking drug; 1. Taking drug<br>Q2 0. No; 1. Yes, at least one | |

| | | |
|---|---|---|
| **Brazil** | Measured in non-fasting serum (mg/dL) (Z031)<br>1. Have you ever been diagnosed with high cholesterol? (Q060)<br>2. Has any doctor or other health care professional given you any recommendations to take medication for high cholesterol? (Q6204) | Z031=total_chol_measure<br>Unless otherwise stated, all questions were answered as: 1. Yes; 2. No |

***Glycaemia:*** To assess CVH, both the LS7 and LE8 criteria recommend fasting plasma glucose, with LE8 additionally providing guidance to score either fasting or non-fasting glycated haemoglobin (HbA1c)[11,12]. In Bangladesh glucose was analysed in fasting serum samples using an autoanalyser (Selectra Pro M). Ethiopia measured fasting blood glucose in capillary samples using a CardioCheck PA Analyser. NHANES measured glucose in fasting plasma samples using a Roche/Hitachi Cobas C Chemistry Analyzer-C311. Both HSE and PNS measured HbA1c in non-fasting plasma samples using a Tosoh G8 Glycohemoglobin Analyser in HSE and high-performance liquid chromatography in Brazil. Current self-reported medication use for diabetes was assessed in all surveys. Harmonization was undertaken using either fasting blood glucose (as capillary and venous glucose levels are reported as more similar in the fasting state[17,18]) or HbA1c values.

| | **Glycaemia** | **(glycemia_measure) Glucose or HbA1c depending on country dataset** | **Glucose:**<br>0. ≥126 mg/dL (treated/untreated); 1. <126 mg/dl (treated); 1. 100-125 mg/dL (untreated); 2. <100 mg/dL (untreated)<br>**Glycated haemoglobin:**<br>0. ≥6.5% (treated/untreated); 1. <6.5% (treated); 1. 5.7 – 6.4% (untreated); 2. <5.7% (untreated) |
|---|---|---|---|
| **Bangladesh** | Measured in fasting serum (glucose mg/dL) (b5)<br>1. Was your blood glucose measured? (h6)<br>2. Were you told you have/had high glucose/diabetes? (h7a)<br>3. Were you told in the past 12 months you have high glucose? (h7b)<br>4. Have you taken meds for high glucose prescribed by a doctor/health worker? (hx5)<br>5. Have you taken meds for high glucose in the past 2 weeks? (h8) | b5=glycemia_measure<br>Unless otherwise stated, all questions were answered as: 1. Yes; 2. No | curr_tx_gluc=Current medication usage for diabetes (1=YES, 2=NO; **Suppl. Figure 1**)<br><br>**For glucose:**<br>Recoded as categorical variable:<br>IF (glycemia_measure >125) gluc=0.<br>DO IF (curr_tx_gluc=1).<br>IF (glycemia_measure <126) gluc=1.<br>END IF.<br>DO IF (curr_tx_gluc=2). |
| **Ethiopia** | Measured in fasting serum (glucose mg/dL) (b5)<br>1. Was your blood glucose measured? (h6)<br>2. Were you told you have/had high glucose/diabetes? (h7a) | | |

<table>
<tr><td></td><td>3. In the past two weeks, have you taken any drugs (medication) for diabetes (prescribed)? (h8)</td><td></td><td rowspan="4">IF (glycemia_measure >99) AND (glycemia_measure <126) gluc=1.<br>IF (glycemia_measure<100) gluc=2.<br>END IF.<br>For HbA1c:<br>IF (glycemia_measure>6.4) gluc=0.<br>DO IF (curr_tx_gluc=1).<br>IF (glycemia_measure <6.5) gluc=1.<br>END IF.<br>DO IF (curr_tx_gluc=2).<br>IF (glycemia_measure >5.6) AND (glycemia_measure <6.5) gluc=1.<br>IF (glycemia_measure<5.7) gluc=2.<br>END IF.</td></tr>
<tr><td>US</td><td>Measured in fasting plasma (glucose mg/dL (LBXGH)<br>1. Were you told you have diabetes? (DIQ010)<br>2. Are you currently taking insulin? (DIQ050)</td><td>LBXGH=glycemia_measure<br>Unless otherwise stated, all questions were answered as: 1. Yes; 2. No</td></tr>
<tr><td>England</td><td>Measured in non-fasting whole blood (HbA1c %) (glyhbval2)<br>1. Have you been diagnosed with diabetes (excluding pregnancy)? (diabete2)<br>2. Do you currently inject insulin for diabetes? (Insulin)<br>3. Are you currently taking any medicines, tablets, or pills for diabetes? (DiMed)</td><td>glyhbval2=glycemia_measure<br>Unless otherwise stated, all questions were answered as: 1. Yes; 2. No</td></tr>
<tr><td>Brazil</td><td>Measured in non-fasting whole blood (HbA1c %) (Z034)<br>1. Have you ever been diagnosed with high glucose/diabetes? (Q030)<br>2. In the past two weeks, because of diabetes, have you taken oral medications to lower your blood sugar? (Q03401)</td><td>Z034=glycemia_measure<br>Unless otherwise stated, all questions were answered as: 1. Yes; 2. No</td></tr>
<tr><td colspan="4">Dietary Quality: To assess diet quality for the CVH score, the 2010 score recommended assessing the intake of five dietary components – fruit and vegetables, fish, fiber-rich wholegrains, sodium and sugar-sweetened beverages.[12] The updated score requires data on both the quantity and types of fruits and vegetables, proteins, fats and grains consumed and on the amount of sodium, added sugar and dairy consumed, with recommendations made for two dietary assessment instruments that can be used to collect this information.[11] Bangladesh and Ethiopia included questions on salt and oil use and the frequency of eating fruit and vegetables, and eating outside of the home, with Bangladesh further asking about frequency of snacking on locally consumed snack foods. Brazil used a food consumption questionnaire composed of 22 locally relevant food items.[19] The US NHANES used a 24-hour dietary recall interview with additional questions to establish if the 24-hour period assessed was usual, alongside typical salt use behaviour and seafood consumption to generate nutrient intakes.[20] The HSE included questions on fruit and vegetable consumption and analysed sodium and potassium in urine.[9] None of these surveys used the diet assessment instruments recommended by AHA for CVH scoring, with differences between countries in data availability, collection methods, and outcomes precluding comparability. Therefore, the decision was made to exclude dietary data as a CVH metric.</td></tr>
<tr><td></td><td>Healthy diet score</td><td>Not possible to harmonize due to lack of data</td><td></td></tr>
<tr><td>Bangladesh</td><td>Components available: Fruit and vegetables, Salt intake</td><td></td><td></td></tr>
<tr><td>Ethiopia</td><td>Components available: Fruit and vegetables, Salt intake</td><td></td><td></td></tr>
<tr><td>US</td><td>Components available: Salt intake, Fish consumption</td><td></td><td></td></tr>
<tr><td>England</td><td>Components available: Fruit and vegetables, Sugar-sweetened beverages</td><td></td><td></td></tr>
</table>

| | |
|---|---|
| | Nuts, legumes, and seeds (pulses) |
| **Brazil** | *Components available:* Meat consumption (chicken and red meat) |

***Sleep:*** The updated CVH score requires estimating average hours of sleep per night. Both Brazil and Ethiopia asked about frequency of sleep problems in the last two weeks as part of the nine item Patient Health Questionnaire to screen for depression.[21 22] Bangladesh and England included no questions on sleep. Only NHANES recorded data on usual sleep and wake times to calculate the average hours of sleep per night.[23] As three of the five surveys did not have the required data, this CVH metric was excluded.

| | **Healthy sleep** | **Not possible to harmonize due to lack of data** |
|---|---|---|
| **Bangladesh** | Is your sleep often interrupted? | |
| **Ethiopia** | Do you have trouble falling asleep, staying asleep or sleeping too much? | |
| **US** | 1. What is your usual sleep time (weekdays/workdays)?<br>2. What is your usual wake time (weekdays/workdays)?<br>3. What is your usual sleep time (weekends)?<br>4. What is your usual wake time (weekends)?<br>5. How many hours do you sleep on weekends?<br>6. How often do you snort or stop breathing?<br>7. Have you ever told a doctor you have trouble sleeping?<br>8. How often do you oversleep during the day? | |
| **England** | *Data not available* | |
| **Brazil** | *Data not available* | |

COVARIATES

| COUNTRY | QUESTION (variable name) | VALUE | CODE ASSIGNED |
|---|---|---|---|
| | **DEMOGRAPHIC VARIABLES** | | |
| | **Age** | **(age_cat)** | 1. 18-24; 2. 25-34; 3. 35-44; 4. 45-54; 5. 55-64; 6. 65-69 |
| **Bangladesh** | How old are you? (c3) | Years (no data with decimal places) | Recoded as:<br>IF (c3>17<25) age_cat=1.<br>IF (c3>24<35) age_cat=2.<br>IF (c3>34<45) age_cat=3.<br>IF (c3>44<55) age_cat=4.<br>IF (c3>54<65) age_cat=5.<br>IF (c3>64<70) age_cat=6. |
| **Ethiopia** | How old are you? (c3) | | |
| **US** | What is your birthdate? (Calculated age from birthdate RIDAGEYR) | | Recoded as:<br>IF (RIDAGEYR>17<25) age_cat=1.<br>IF (>24<35) age_cat=2.<br>IF (RIDAGEYR>34<45) age_cat=3.<br>IF (RIDAGEYR>44<55) age_cat=4.<br>IF (RIDAGEYR>54<65) age_cat=5.<br>IF (RIDAGEYR>64<70) age_cat=6. |
| **England** | What is your date of birth? (Calculated age from birthdate and categorized to within 5-year age bands Age16g5)<br>1.16-17 yrs<br>2.18-19 yrs<br>3.20-24 yrs<br>4.25-29 yrs<br>5.30-34 yrs<br>6.35-39 yrs<br>7.40-44 yrs<br>8.45-49 yrs<br>9.50-54 yrs<br>10.55-59 yrs<br>11.60-64 yrs<br>12.65-69 yrs<br>13.70-74 yrs<br>14.75-79 yrs | | Recoded as:<br>IF (Age16g5=2 OR Age16g5=3) age_cat=1.<br>IF (Age16g5=4 OR Age16g5=5) age_cat=2.<br>IF (Age16g5=6 OR Age16g5=7) age_cat=3.<br>IF (Age16g5=8 OR Age16g5=9) age_cat=4.<br>IF (Age16g5=10 OR Age16g5=11) age_cat=5.<br>IF (Age16g5=12) age_cat=6. |

| | | | |
|---|---|---|---|
| | 15.80-84 yrs<br>16.85-89 yrs<br>17.90+ | | |
| **Brazil** | How old are you? (Z002) | | Recoded as:<br>IF (Z002>17<25) age_cat=1.<br>IF (Z002>24<35) age_cat=2.<br>IF (Z002>34<45) age_cat=3.<br>IF (Z002>44<55) age_cat=4.<br>IF (Z002>54<65) age_cat=5.<br>IF (Z002>64<70) age_cat=6. |
| | **Sex/Gender** | **(sex)** | 0. Women; 1. Men |
| **Bangladesh** | Observed (c1) | Unless otherwise stated, sex/gender was answered as:<br>1. Male/men<br>2. Female/women | Recoded as:<br>2 → 0<br>1 → 1 |
| **Ethiopia** | Observed (c1) | | |
| **US** | Ask if not obvious to observe (RIAGENDR) | | |
| **England** | Observed (Sex) | | |
| **Brazil** | Observed (Z001) | | |
| | **Education** | **(education)** | 0. Primary school or less; 1. Secondary / High school; 2. College / University degree; 3. Foreign |
| **Bangladesh** | What is the highest level of education you have completed? (c5) | 1. No formal schooling<br>2. Less than primary school<br>3. Primary school completed<br>4. Secondary school completed<br>5. High school completed<br>6. College / University completed<br>7. Post graduate degree | Recoded as:<br>IF (c5=1 OR c5=2 OR OR c5=3) education=0.<br>IF (c5=4 OR c5=5) education=1.<br>IF (c5=6 OR c5=7) education=2. |
| **Ethiopia** | What is the highest level of education you have completed? (c5) | 1. No formal schooling<br>2. Less than primary school<br>3. Primary school completed<br>4. Secondary school completed<br>5. College / University completed<br>6. Post graduate degree | |
| **US** | What is the highest grade or level of school you have completed or the highest degree you have received? (DMDEDUC2) | 1. Less than $9^{th}$ grade<br>2. $9^{th}$ to $11^{th}$ grade (including $12^{th}$ grade with no diploma) | Recoded as:<br>IF (DMDEDUC2=7 OR DMDEDUC2=9 OR DMDEDUC2=1) education=0.<br>IF (DMDEDUC2=2 OR DMDEDUC2=3) education=1. |

| | | | |
|---|---|---|---|
| | | 3. High school graduate / GED or equivalent<br>4. Some college or AA degree<br>5. College graduate or above<br>9. Don't know | IF (DMDEDUC2=4 OR DMDEDUC2=5) education=2. |
| **England** | Which of the qualifications on this card do you have? (topqual3) | 1. NVQ4/NVQ5/Degree or equivalent<br>2. Higher ed below degree<br>3. NVQ3/GCE A Level equivalent<br>4. NVQ2/GCE O Level equivalent<br>5. NVQ1/CSE other grade equivalent<br>6. Foreign/other<br>7. No qualification | Recoded as:<br>IF (topqual3=4 OR topqual3=5) education=1.<br>IF (topqual3=1 OR topqual3=2 OR topqual3=3) education=2.<br>IF (topqual3=6) education=4. |
| **Brazil** | What is the highest level of education achieved? (VDD004) | 1. No instruction<br>2. Incomplete elementary or equivalent<br>3. Complete fundamental or equivalent<br>4. Incomplete high school or equivalent<br>5. Complete high school or equivalent<br>6. Incomplete University degree or equivalent<br>7. University degree and above | Recoded as:<br>IF (VDD004=1 OR VDD004=2 OR VDD004=3) education=0.<br>IF (VDD004=4 OR VDD004=5) education=1.<br>IF (VDD004=6 OR VDD004=7) education=2. |
| | **Rural/Urban residence – Data not available for harmonization** | | |
| **Bangladesh** | Locality of the respondents (urbanrural) | 1. Urban<br>2. Rural | |
| **Ethiopia** | Locality of the respondents (urbanrural) | 1. Urban<br>2. Rural | |
| **US** | *Data not available* | | |
| **England** | Rurality of dwelling unit (urban/rural) (Urban14b) | 1. Urban<br>2. Town/ Fringe/ Village, hamlet, and isolated dwellings | |
| **Brazil** | *Data not available* | | |
